# Using transportability methods to map the local effectiveness of mass drug administration for malaria in Senegal

**DOI:** 10.1101/2025.08.09.25333369

**Authors:** Michelle E. Roh, Yanwei Tong, Gabriella Barratt Heitmann, El-hadji Konko Ciré Ba, Jean Louis Ndiaye, Ari Fogelson, Paul Milligan, Amadou Seck, Abdoulaye Diallo, Aminata Colle Lo, Michael Baiocchi, Roly Gosling, Adam Bennett, Michelle S. Hsiang, Jade Benjamin-Chung

**Author notes:** **Corresponding author:** Jade Benjamin-Chung, PhD, 300 Pasteur Drive, MC: 4505, Stanford, CA, USA 94305.

## Abstract

Numerous trials have evaluated the effectiveness of mass drug administration (MDA) to rapidly reduce malaria transmission, but it is unknown whether estimated effects generalize to other populations eligible for MDA. A recent cluster-randomized trial in Senegal found that MDA reduced malaria incidence by 55% in areas routinely deploying seasonal malaria chemoprevention (SMC). Here, we used transportability models with machine learning to generalize trial effects to 116 non-trial Communes where SMC is standard-of-care. Accounting for differences in weather, vegetation, and population density between trial and non-trial areas, we estimated significant reductions in incidence of 36%–65% in 74 non-trial Communes, with larger reductions in areas with higher precipitation, denser vegetation, and lower temperatures. We found that MDA was not effective in the post-intervention year in non-trial Communes, supporting the notion that MDA effects are short-lived. Our approach offers a scalable framework for generalizing trial findings to target environmentally-mediated infectious disease interventions.

## Introduction

Malaria remains a major global public health burden, with an estimated 263 million cases and 597 000 attributable deaths in 2023.^1^ Despite significant advances in reducing the disease burden, progress towards malaria elimination has stalled. This stagnation has been particularly pronounced in countries with moderate-to-high transmission,^1^ where more intensive or novel strategies may be required to accelerate progress.

Mass drug administration (MDA) has regained attention as a potential strategy to accelerate toward malaria elimination. MDA involves administering a full treatment course of antimalarials to all individuals within a defined geographic area, regardless of infection status, with the frequency and duration of rounds tailored to its goal (i.e., to either rapidly reduce burden or interrupt transmission). To date, 12 cluster-randomized controlled trials have evaluated MDA for transmission reduction,^2–4^ including 4 from moderate-to-high transmission settings.^4,3,5,6^ A recent meta-analysis of 10 of these trials^2^ reported mixed findings of the effectiveness of MDA, with most reporting only short-term reductions in incidence and/or prevalence in very low-to-low transmission settings.^2–4^ Consistent with these findings, mathematical models also suggest the effects of MDA are likely time-limited and highly context-dependent.^7^ As such, the World Health Organization (WHO) recommends MDA for very low-to-low transmission settings, but not for moderate or high transmission settings due low certainty of evidence.^8^

Although numerous cluster-randomized trials and meta-analyses have evaluated the effectiveness of MDA,^2,9^ the generalizability of these results to broader settings remains unclear. While cluster-randomized trials are inherently designed to yield internally valid estimates (by minimizing confounding through randomization), they may offer limited external validity for two key reasons. First, cluster-randomized trials typically estimate treatment effects as average differences between intervention and control clusters. This approach may obscure meaningful heterogeneity in intervention impact, especially when contextual factors such as transmission intensity, vector ecology, or human mobility vary across clusters.^10–12^ Second, cluster-randomized trials often include a limited number of clusters and impose strict eligibility criteria to facilitate operational feasibility, enrollment, statistical power, intervention delivery, and follow-up,^13^ which can constrain generalizability. These limitations carry forward in meta-analyses, which often pool data from trials to increase statistical power. However, most trials are conducted in a limited number of settings and may not represent the target populations to which interventions would be applied. As a result, the pooled effect estimates, which form the basis for WHO policies, can also have low external validity. Because MDA effectiveness may be highly context-dependent, understanding drivers of heterogeneity in MDA effectiveness is critical to subnational targeting to maximize its cost-effectiveness, a priority outlined by the WHO Global Technical Strategy for Malaria 2016–2030.^14^

In recent years, transportability models have been developed in the causal inference literature to quantitatively assess the external validity of studies by extrapolating intervention effect estimates from trials to external target populations.^15^ These models require data on factors that modify the intervention effect (i.e., “effect modifiers”) and differ in distribution between trial and target populations.^16^ A key advantage of this approach is that it does not strictly require outcome data (e.g., malaria incidence or prevalence) from the external target population. Instead, transportability models use information on the distribution of effect modifiers in both study and target (non-trial) populations to estimate intervention effects beyond the original study setting. For environmentally-mediated infections like malaria, fine-scale remote sensing data has immense potential as a source of effect modifier data in transportability analyses. Moreover, remote sensing data are widely available, typically free of charge, cover large geographic areas, and offer high spatial and temporal resolution making them well suited for transporting intervention effects across entire countries and regions.

Transportability models are complementary to mathematical models, which the WHO recommends for informing data-driven subnational targeting of interventions.^14^ These models integrate mechanistic understanding of transmission dynamics to simulate the potential impact of interventions under various scenarios. However, they often rely on strong assumptions and parameter inputs that are difficult to validate and may not accurately reflect local conditions.^17^ Further, competing models often produce substantially different intervention effect estimates.^7^ As a result, findings might not hold in populations with differing epidemiologic or entomological characteristics.^18^ On the other hand, transportability analyses leverage real-world trial data and localized effect modifier data to estimate intervention effects in new populations, thereby providing an additional framework to inform targeting and tailoring of interventions for maximal benefit.

Here, we transport effects of MDA estimated from a recent cluster-randomized trial in Senegal^4^ to non-trial areas of Senegal where SMC is offered as standard-of-care. Our goal was to characterize geographic areas where MDA may be more or less effective, accounting for differences in environmental and demographic variables.

## Results

### Original trial results

We conducted a secondary analysis of a two-arm, cluster-randomized controlled trial evaluating the impact of MDA on malaria transmission (NCT04864444).^4^ The trial was conducted from September 2020 to December 2022 in a low-to-moderate transmission setting of southeastern Senegal. Sixty villages were assigned to receive intervention or control using stratified, constrained randomization. In intervention villages, individuals aged ≥3 months received three rounds of MDA with dihydroartemisinin-piperaquine plus single low-dose primaquine every six weeks, initiated on June 21, 2021, approximately one month before the start of the transmission season (July–December) to clear the parasite reservoir. In control villages, children 3-120 months received standard-of-care SMC with sulfadoxine-pyrimethamine plus amodiaquine every four weeks at the presumed start of the transmission season (July 30, 2021). MDA was implemented for one year (2021), after which all villages resumed SMC in 2022. Before the intervention, all villages received pyrethroid-piperonyl butoxide (PBO) bednets at high coverage (∼98%).^4^ The timeline of study activities is shown in **Figure 1**.

**Figure 1.**
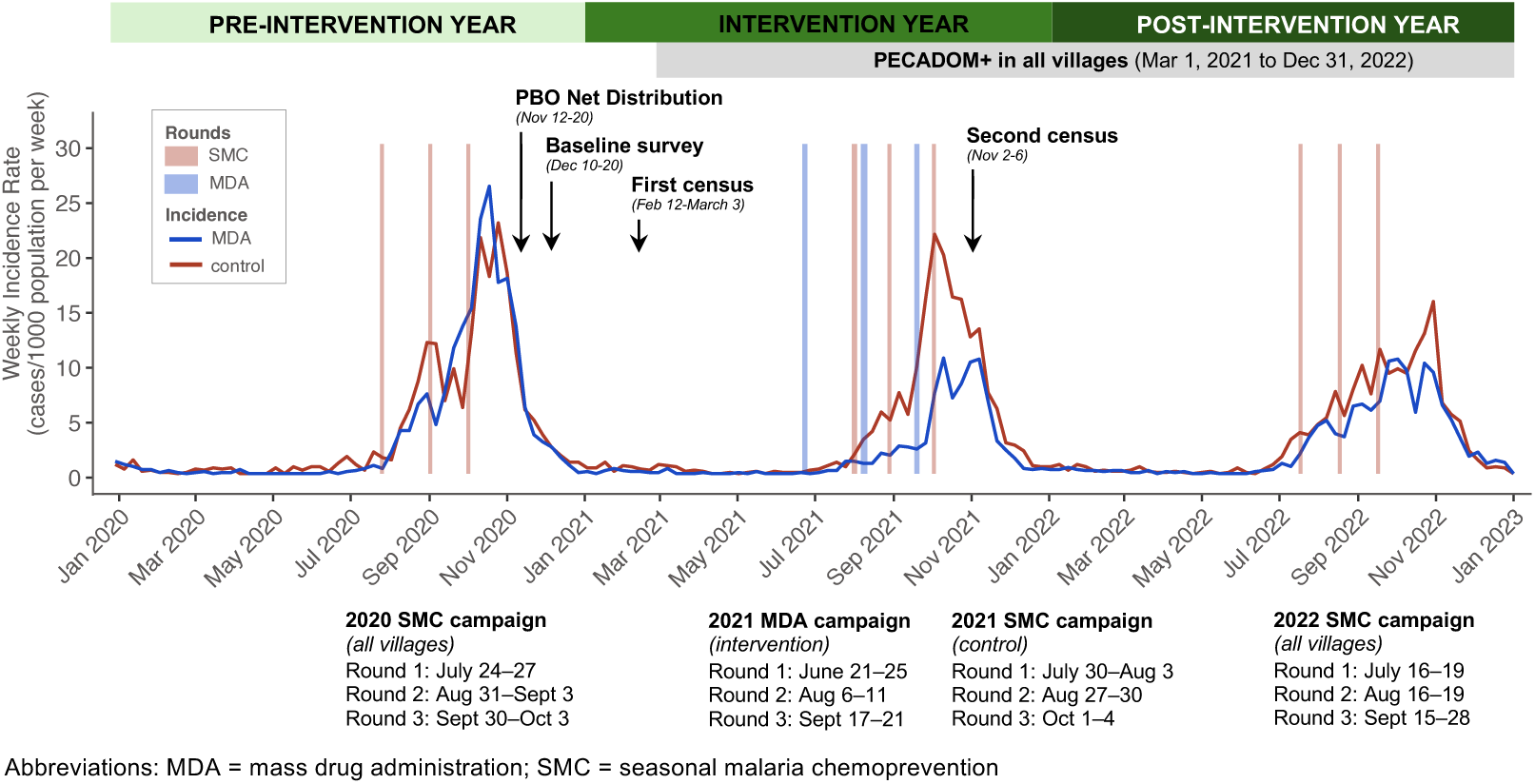
Trial activities and epidemiological curve of weekly malaria incidence in the pre-intervention, intervention, and post-intervention years.

During the intervention year (2021), environmental variables and travel time to the nearest health facility were well-balanced between arms (**Appendix Table 1**). Population density was higher in control villages compared to MDA. Median village-level coverage was 68%, 73%, and 74% across the three MDA rounds and 93%, 92%, and 92% across the three SMC rounds. In the original trial, MDA reduced clinical malaria incidence by 55% (95% CI: 28, 71) during the transmission season of the intervention year (July–December 2021) and by 26% (95% CI: −17, 53) in the following transmission season (July–December 2022), when all villages resumed SMC.^4^

### Effect heterogeneity within trial clusters

To identify candidate effect modifiers for transportability analyses, we assessed whether MDA effects varied by spatial, environmental, and demographic factors (**Figure 2**). Effect modification was assessed on both the multiplicative and additive scale (**Appendix Tables 2-4**).^19^ Spatial variation was assessed by estimating effects at the Commune-level—the smallest standard administrative level used for public planning in Senegal and the scale at which trial estimates were transported. Environmental variables were lagged 1-3 months to account for delayed impacts on vector breeding, survival, and biting.

**Figure 2.**
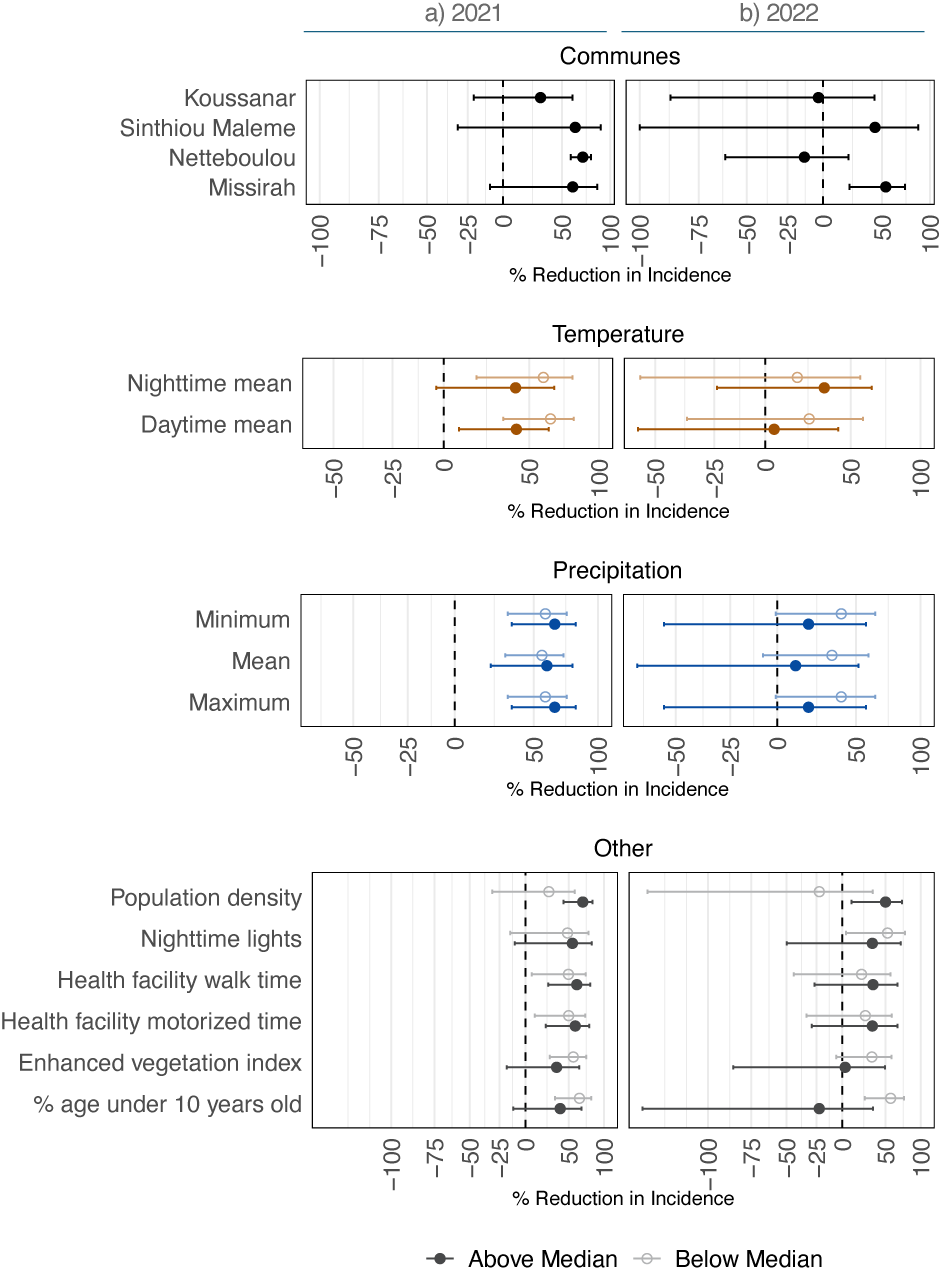
Heterogeneous effects of mass drug administration in the intervention and post-intervention years. Point estimates represent adjusted incidence ratios from subgroup analyses comparing the impact of MDA on malaria incidence during the intervention year (a) and the post-intervention year (b) across potential spatial, environmental, and demographic effect modifiers. For commune subgroup analyses, incidence ratios compare incidence in the MDA arm to the control arm. Other effect modifiers were coded as binary indicators for values above or below the median in a given year. Analyses were performed at the cluster level; analysis dataset included pre-intervention, intervention year, and post-intervention years (N=3 years) and 60 clusters (30 per arm) (N=180). Horizontal bars indicate 95% confidence intervals computed with robust standard errors. In (b), the lower bound for Sinthiou Maleme was imputed for clarity of visualization; the estimated value was −104%. Incidence ratios were adjusted for timing of case detection relative to full PECADOM+ scale-up, similar to the original trial analysis

Across the four Communes that encompassed trial villages (Koussanar, Sinthiou Maleme, Netteboulou, and Missirah), we observed substantial heterogeneity in MDA effects (**Figure 2; Appendix Table 2**). During the intervention year (2021), effect sizes in the three southernmost Communes (Sinthiou Malame, Netteboulou, and Missirah) were greater than the overall effect estimate in the original trial (% reduction: 55% (95% CI: 28, 71)^4^). In Netteboulou, the percent reduction in malaria incidence was 70% (95% CI: 58, 79). In Sinthiou Maleme and Missirah, the estimated impact of MDA was a reduction of approximately 60%, although 95% CIs included the null. In the post-intervention year (2022), MDA did not have a statistically significant impact on incidence (similar to the original trial estimate), except in the southernmost Commune of Missirah (54% (95% CI: 21, 73) vs. 26% (95% CI: −17, 53)^4^). Tests of interaction between Commune and treatment arm did not reach the 0.05 threshold for statistical significance on either the ratio or additive scales in 2021 (**Appendix Table 3**). In 2022, there was evidence of interaction on the ratio scale in Netteboulou and Missirah and on the additive scale in Missirah (**Appendix Table 4**).

Next, we linked village centroids to publicly available remote sensing data to examine heterogeneity in intervention effects by temperature, precipitation, enhanced vegetation index (EVI), population density, nighttime lights, and travel time to nearest health facility. These variables were selected a priori based on the hypothesis that they may modify MDA effectiveness: environmental conditions are known to influence malaria transmission dynamics, travel time to health facility may serve as a proxy for MDA coverage in real-world implementation settings, and nighttime light radiance may reflect underlying household socioeconomic status. For each variable, we conducted stratified analyses based on cluster values that fell above or below median (**Figure 2, Appendix Table 2**). During the intervention year, MDA was more effective in areas with below-median mean temperature levels (additive scale interaction p-values = 0.072 for mean daytime temperature, 0.107 for mean nighttime temperature) (**Appendix Table 3**). In both the intervention and post-intervention years, MDA was more effective in areas with above-median population density (ratio scale interaction p-value = 0.020 in 2021, 0.091 in 2022), and below-median proportion of the population aged under 10 years (ratio scale interaction p-value = 0.048 in 2021, 0.008 in 2022) (**Appendix Tables 3-4).**

### Heterogeneity of environmental and demographic characteristics across Senegal

To assess the potential for transporting trial findings across Senegal, we examined the spatial heterogeneity of potential effect modifiers at the national level. For each Commune, we mapped mean values during the 2021 and 2022 transmission seasons (**Figure 3**). Communes where SMC was routinely implemented had lower mean temperatures, nighttime light radiance, and population density, and higher total precipitation, and EVI compared to non-SMC implementing Communes. Within SMC-implementing areas, population density exhibited the greatest spatial variability between Communes (coefficient of variation (CV)=109); CVs for motorized time to nearest health facility, nighttime lights, walking time to nearest health facility, mean precipitation, EVI, daytime temperature, and the proportion of the population <10 years, and were 70, 67, 65, 26, 18, 9, 6, respectively. The extent to which values of effect modifiers in the trial site were similar to non-trial sites varied by follow-up month. For example, EVI and precipitation in the trial site was higher than in most non-trial sites from July–September and lower in other months (**Appendix Figure 1**).

**Figure 3.**
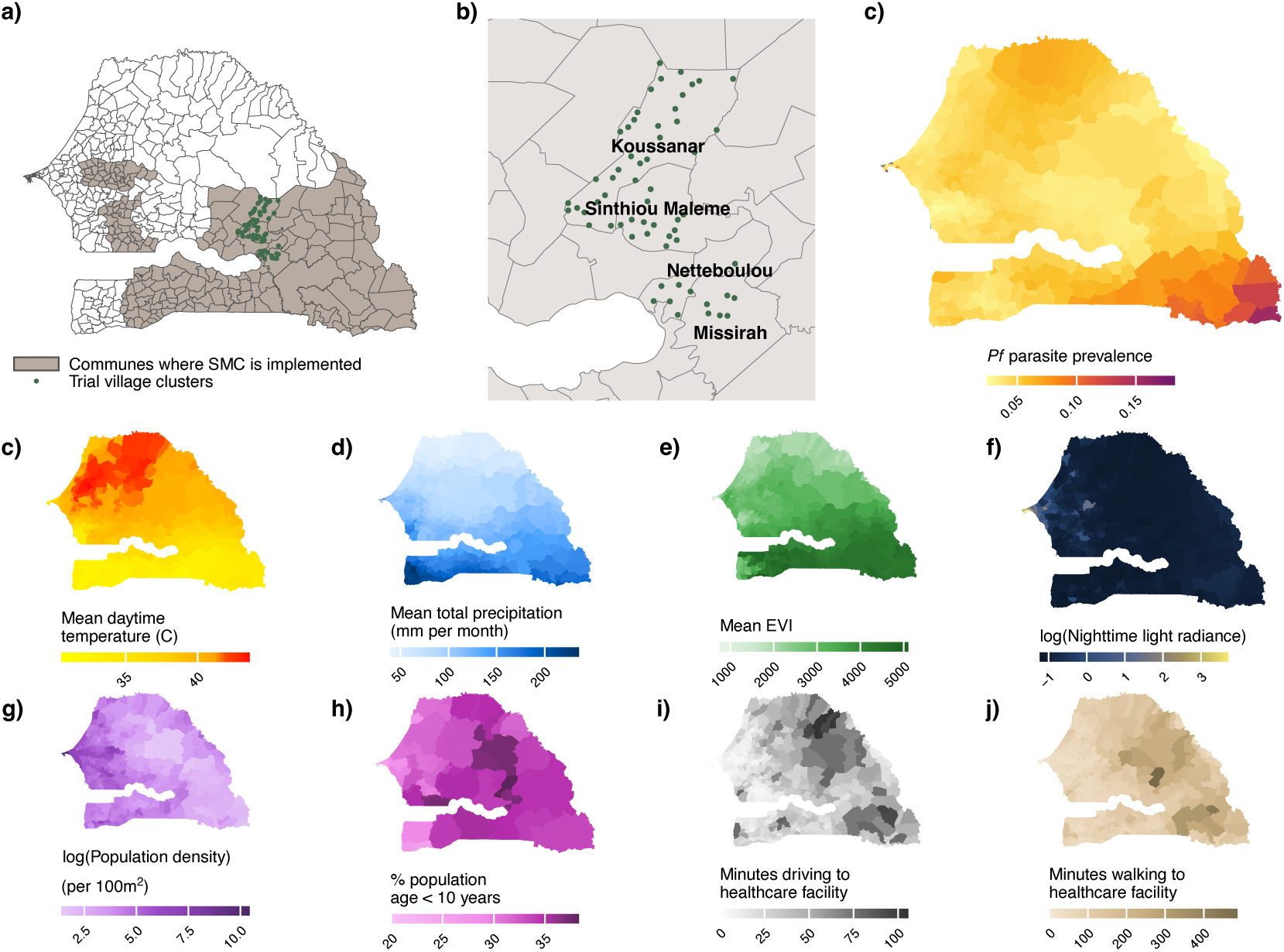
Maps depicting areas routinely implementing seasonal malaria chemoprevention (SMC), location of trial villages, and variation in environmental and demographic covariates. In Panel (a), grey shaded areas indicate where seasonal malaria chemoprevention was routinely implemented in Senegal during the trial period; green points indicate the 60 clusters included in the original trial. Panel (b) depicts trial villages (green points) nested within the 4 Communes encompassing the study site. Panel (c) depicts predicted *Plasmodium falciparum* malaria prevalence in 2021-2022 extracted from the Malaria Atlas Project. Panels (c-h) depicts values of environmental and demographic variables across Senegal used to assess effect heterogeneity. Panels (i-j) depict values of travel time (in minutes) to the nearest health facility. Values are shown as the mean value in each commune from July-December transmission seasons in 2021 and 2022.

### Transporting MDA effects to non-trial areas

To estimate the impact of MDA in areas beyond the trial’s geographic footprint, we conducted transportability analyses using a doubly robust modeling approach.^20^ This method treats trial clusters as a sample of a broader target population, defined here as Communes in Senegal where SMC is implemented as standard-of-care. As the trial was conducted under conditions of high vector control coverage, prompt malaria case management, and robust surveillance, transported effect estimates assumed comparable programmatic conditions between trial and non-trial areas.

Of the 433 Communes in Senegal, 129 were eligible for transportability analyses. Of these, 4 Communes (Koussanar, Sinthiou Maleme, Netteboulou, Missirah) included trial villages. Communes were excluded if SMC was not implemented as standard of-care (n=247) and/or population density exceeded that of the trial site (>152 people per 100 m^2^) (n=57) (**Appendix Figure 2**). The distributions of most potential effect modifiers overlapped between the trial clusters and the 129 eligible non-trial Communes (**Appendix Figure 3**). The effect modifiers included in transportability models were precipitation, temperature, EVI, and population age distribution (see details in **Methods**).

Our doubly robust modeling approach relied on two components. First, for each Commune, we fit a model to estimate the probability that the Commune would have been included in the trial at month *t* based on their effect modifier values. The predicted probabilities for each Commune were skewed and right-modal, suggesting Communes were likely to be representative of the target population when accounting for differences in effect modifiers (**Appendix Figure 4**). There was substantial overlap in the probabilities in trial and non-trial areas for most Communes, which is required for causal identification in transportability analyses. Next, we modeled expected malaria incidence in each Commune at month *t* as a function of effect modifiers using trial data, then predicted outcomes in each arm in non-trial Communes. We then implemented a doubly robust estimator that combines both approaches and is consistent if either the outcome or selection model is correctly specified.^20^ For each SMC-implementing Commune, we estimated MDA effectiveness as (1 - incidence ratio) x 100%. We performed separate transportability analyses for the intervention and post-intervention years. Outcome models used for transportability did not account for baseline malaria incidence or include a population offset, as was done in the original trial because such data was not available in non-trial areas. Comparison of estimates from transportability models and the original trial showed minimal differences in point estimates and 95% CIs in the intervention year and moderate differences in the post-intervention year, though interpretation of findings was similar between approaches with overlapping 95% CIs (**Appendix Tables 5–6**).

### Estimated effects of MDA in non-trial areas

Of 129 eligible Communes, we were able to transport estimates for 116 during the intervention year (**Figure 4a**) and 118 in the post-intervention year (**Appendix Figure 5a**). Estimates were not transported in 13 Communes in the intervention year and 11 Communes in the post-intervention year due to potential positivity violations (i.e., probability of trial participation <0.75). During the intervention year, the predicted MDA effectiveness varied across Communes, ranging from −23–65% (**Figures 4–5**). In 74 (64%) of Communes, we estimated statistically significant reductions that ranged from 36–65%; with estimates exceeding the original trial estimate in 37 (32%) Communes (range: 56–65%). The effect size and precision of estimates, defined by the width of 95% CIs, was generally higher in Communes located in the southeastern region of Senegal, where environmental and demographic characteristics were more similar to the trial site (**Figure 4c**). In the post-intervention year, most transported effects were closer to the null compared to the original trial estimate (26% (95% CI −18%, 53%)^4^), with none reaching statistical significance (**Appendix Figure 5**).

**Figure 4.**
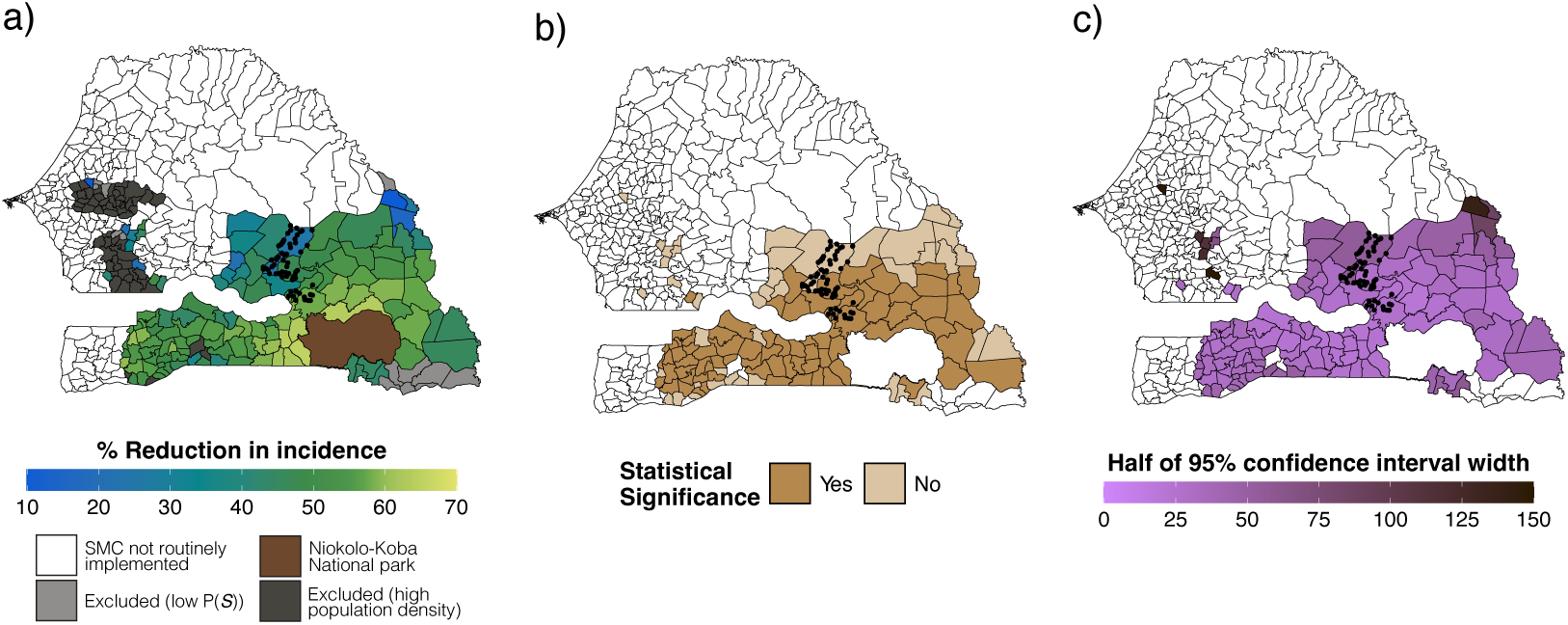
Spatial distribution of transported effects of mass drug administration. Polygons indicate communes. Panel (a) shows the percent reduction in malaria (1 – the ratio of cases in the MDA vs. control arm) during the transmission season of the intervention year (2021), estimated using doubly robust transportability models. Covariates included precipitation, temperature, enhanced vegetation index, population density, and % of population <10 years. The final covariate list included in final models varied for each Commune-month following screening for collinearity, data sparsity, association with the malaria case count, and feature selection using elastic net regression. Transportability analyses were restricted to communes where SMC was routinely offered during the trial period, had a population size <152 per km, and a predicted probability of trial selection (P(*S*)) of >0.75. Separate analyses were performed for each Commune-month (N=366 per Commune). Panel (b) indicates whether the 95% confidence intervals for transported estimates included the null. Panel (c) shows uncertainty, depicted as half the length of the 95% confidence interval, estimated using a non-parametric bootstrapping procedure that resampled commune-months within trial datasets and months within the non-trial Commune 1,000 times with replacement.

**Figure 5.**
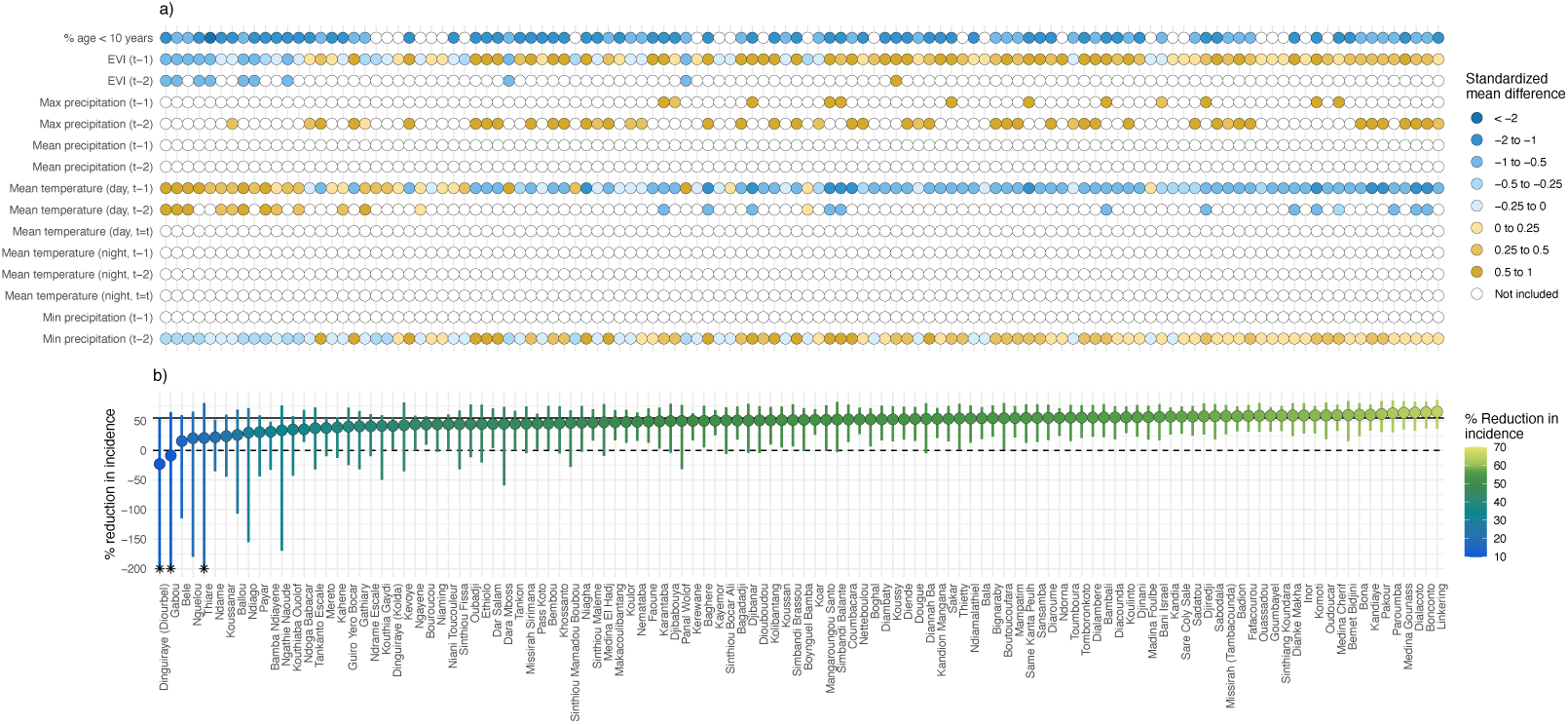
Transported effect estimates of mass drug administration to non-trial areas during the intervention year and covariates included in transportability analysis. Panel (a) depicts covariates used in the transportability analysis for each commune. Colored points indicate covariates included in the transportability models after screening for collinearity, data sparsity, association with the malaria case count, and feature selection using elastic net regression. Blue and yellow shaded points indicate the standardized mean difference for a given covariate in a given commune, calculated as the mean in a non-trial commune minus the mean in the trial site divided by the pooled standard deviation. White points indicate covariates that did not pass screening. In Panel (b), colored points indicate the transported effect for each Commune, expressed as the percent reduction in malaria incidence (1 – the ratio of cases in the MDA vs. control arm) during the transmission season of the intervention year, using doubly robust transportability models. Analyses were restricted to communes with population size σ152 per km, a predicted probability of trial participation >0.75, and where SMC was offered during the trial period. Separate analyses were performed for each commune using monthly data from July to December (N=366 per commune). Analyses used doubly robust transportability models. Vertical colored lines indicate 95% confidence interval for each commune, which was obtained from non-parametric bootstrap that resampled commune-months 1,000 times with replacement. Communes are sorted by transported effect size. The grey solid like indicates the original trial estimate (55% (95% CI: 28, 71). Asterisks indicate confidence intervals that were truncated for data visualization (lower bound < −200% and/or upper bound > 100%).

### Drivers of heterogeneity of transported effects

To explore the potential drivers of heterogeneity in transported effect estimates, we calculated the standardized mean difference for effect modifiers between each Commune and the trial site. During the intervention year, Communes with larger reductions in incidence for MDA vs. control tended to have higher EVI and precipitation levels and lower mean temperatures and % under 10 years compared to Communes with smaller reductions (**Figure 5**). In the post-intervention year, no results reached statistical significance (**Appendix Figure 6**).

### Sensitivity analyses

To validate our transportability model, we transported trial estimates to areas surrounding the trial villages (**Appendix Figure 7**), where effect modifier values should be similar, and thus, transported estimates were expected to align closely with the original trial results. In the intervention year (2021), the transported percent reduction in malaria incidence in the trial site was 50% (95% CI 18%, 71%) compared to 55% (95% CI 28%, 71%) in the original trial and 49% (95% CI: 25%, 66%) using trial data without adjustment for baseline malaria incidence (**Appendix Table 5**).^4^ In the post-intervention year (2022), the transported percent reduction in the trial site was 4% (95% CI −64%, 33%) compared to 26% (95% CI −17%, 53%) in the original trial^4^ and 4% (95% CI −28%, 29%) using trial data without baseline adjustment (**Appendix Table 5**).

Next, because MDA coverage was not uniform across intervention clusters (range: 54–80%), we examined whether this variability was correlated with effect modifiers. If so, it would suggest that our transported effects could, in part, reflect differences in operational factors between study clusters. Correlations were generally weak (|ρ| < 0.2) for most modifiers, except for daytime temperature, precipitation, and travel time to nearest health facility, which demonstrated moderate-to-high correlation with coverage (**Appendix Figure 8**).

## Discussion

In this re-analysis of a cluster randomized trial from Senegal, we generalized trial-based estimates to other parts of the country, with the goal of identifying areas where MDA could be integrated with existing interventions to rapidly reduce malaria burden. Within the trial, we found substantial spatial heterogeneity in the effectiveness of MDA, with the greatest impacts observed in the southernmost Communes. In the intervention year, we estimate that MDA would have significantly reduced malaria incidence by 36–65% in 64% of eligible non-trial Communes. Notably, estimated effects exceeded the original trial’s overall estimate in 32% of Communes during the intervention year, mainly in regions just south of the trial site, which had higher precipitation, denser vegetation, and lower temperatures. However, effects were not sustained upon discontinuation of MDA in any of non-trial Communes, aligning with conclusions from prior studies and WHO recommendations^8^ that MDA effects can have a strong, but transient effect on malaria incidence.

To our knowledge, this is the first study to apply transportability methods to generalize estimates from a large, population-level malaria intervention trial. By integrating trial data with fine-scale remote sensing, this approach offers a powerful, data-driven framework to identify areas where interventions would yield the greatest impact, making it a powerful addition to the existing toolbox for guiding subnational targeting. For MDA and other aggressive malaria interventions, such targeted implementation is critical given its high cost, operational complexity, and heterogeneous effects across epidemiological settings. In this study, 64% of Communes were estimated to benefit from MDA during the intervention year. While these effects were not sustained beyond that period, ongoing trials of more novel interventions (e.g., malaria vaccines; NCT06578572) may offer new opportunities for longer-lasting impact. Transportability methods could play an important role in identifying optimal areas for deployment of these tools.

It is important to note that in the original trial, the number of MDA rounds administered during the intervention year did not sufficiently cover the entire transmission season, limiting its ability to fully clear the infectious reservoir. Monthly incidence data from the original trial supports this notion (**Figure 1**), showing a leveling off of effects in November–December 2021, when MDA rounds were stopped. Thus, it may be possible that additional MDA rounds during these months would have yielded a stronger intervention impact in both the intervention and post-intervention year, thereby potentially increasing the number of Communes that could be targeted for MDA.

This study showcases how transportability models that leverage publicly available remote sensing datasets can be used to extend trial findings beyond a specific study site to inform intervention targeting. While our study likely did not include all potential effect modifiers, we anticipate that weather, temperature, vegetation indices, and the population age distribution accounted for substantial heterogeneity in malaria transmission,^21,22^ as demonstrated by prior studies.^23^ While we explored additional hypothesized modifiers (e.g., predicted *Pf* malaria prevalence, surface water presence, nighttime light radiance, and travel time to nearest health facility), we ultimately excluded these variables due to lack of overlap with trial sites, insufficient variability, or strong collinearity. Thus, we may not have accounted for all potential effect modifiers—a key assumption of transportability models.^15^ One key factor our models did not account for (due to a lack of available data) was healthcare system capacity, which is likely a key operational determinant of MDA coverage and effectiveness in real-world settings.^24^ However, our validation study demonstrated that transported estimates for the area surrounding the trial site were highly similar to original trial estimates, lending credibility to our findings. In future applications, transportability models for malaria interventions could be further strengthened by integrating additional context-specific variables, such as population mobility patterns^25,26^ and local vector species distributions and density. Moreover, use of pooled datasets from multiple trials of the same intervention could provide greater variability across epidemiological settings, enabling more robust transportability analyses of malaria intervention effects over large spatial extents.^27^

This study also had several limitations. First, effect modifier data was aggregated to the month level, which may have masked important daily variation for certain variables such as temperature. Second, we did not explicitly account for coverage of other malaria co-interventions (i.e., vector control and/or diagnostic, case management, and surveillance capacity), which are also likely modifiers of MDA impact. Thus, our transported MDA estimates implicitly assumed that that these co-interventions would be implemented prior to MDA (as recommended by WHO^8^) and at a similar coverage to the trial. Third, the number of study clusters was relatively small (N=60), so we leveraged temporal variability within the follow-up period to fit transportability models. As such, the number of units with combinations of effect modifier values may have been limited in certain strata. Transportability analyses would, in theory, be more robust for trials with a larger number of units and larger geographic footprint. Fourth, our transported estimates do not account for variation in intervention coverage and thus assumes that MDA would have been implemented in non-trial areas at similar level as in the trial and that the relationship between effect modifiers and coverage would hold in transported populations. Fifth, transportability models were unable to account for pre-intervention differences in malaria incidence as in the original trial analysis. In a sensitivity analysis of the original trial analysis without accounting for pre-intervention incidence (**Appendix Table 5**), our point estimates were slightly closer to the null in the intervention year and substantially attenuated towards the null in the post-intervention year. Thus, transported effects for the post-intervention year results should be interpreted with caution as a potentially more conservative, attenuated effect. Finally, we were only able to conduct transportability analyses in a subset of Communes where SMC is implemented in Senegal; other Communes were excluded due to limited overlap in effect modifiers with the trial clusters.

### Conclusion

In summary, our study demonstrates the value of transportability methods as a data-driven tool to guide subnational targeting of interventions. Consistent with existing literature, we estimate that, when implemented with high vector control coverage, prompt case management, and a robust surveillance system, MDA can yield significant short-term reductions in malaria burden across broad geographic contexts, though effects are likely not sustained beyond the intervention period. In today’s malaria prevention landscape, multiple new and highly effective but costly interventions are available (e.g., next-generation insecticide treated nets and malaria vaccines), but funding limitations and other factors make universal rollout infeasible. Our general approach provides a rigorous evidence base for local decisions about subnational intervention tailoring in this changing landscape.

## Methods

### Analysis overview

The present study analyzed data from the Senegal MDA trial (NCT04864444), a cluster randomized controlled trial evaluating MDA for accelerating malaria elimination in low-to-moderate transmission settings of southeastern Senegal. While detailed trial descriptions have been published elsewhere,^4^ we summarize key details relevant to our analyses below.

### Study site

The Senegal MDA trial was conducted in Tambacounda Health District of southeastern Senegal, where malaria transmission is low-to-moderate and highly seasonal, with most cases occurring during the rainy season (July–December). At this study site, the NMP implements standard malaria control measures, including mass distribution of insecticide-treated nets (ITNs), health facility-based malaria case management, and SMC for children aged 3–120 months. In remote areas with limited healthcare access, Senegal employs community malaria case management through the Prise en Charge à Domicile (PECADOM) model, whereby village-level community health workers (dispensateur de soins à domiciles; DSDOMs) are trained to test and treat febrile individuals for malaria using rapid diagnostic tests (RDT) and first-line antimalarials. In some villages, PECADOM has evolved into PECADOM+, a proactive form of the model in which DSDOMs conduct weekly household visits to improve early detection and treatment.

In the rest of the country, malaria endemicity is highly heterogeneous, driven largely by geography, climate, population density and age distribution. To accommodate varying transmission intensities, the NMP adopts a tailored set of interventions according to each transmission stratum. In the northern and central regions, where transmission is very low due to the semi-arid Sahelian climate, the NMP prioritizes pre-elimination activities consisting of case investigation and proactive response measures. In the central and southern regions, where transmission ranges from low to high due to denser vegetation and higher rainfall, the NMP prioritizes malaria control interventions, including routine distribution of ITNs, prompt case management, and SMC in highly seasonal areas. The southeastern districts of Tambacounda, Kolda, and Kédougou bear 81% of Senegal’s malaria burden, with Kédougou having the highest incidence (>400 cases/1000 population).

### Study design

The Senegal MDA trial utilized a two-arm, open-label cluster randomized controlled trial design. Sixty villages were randomly selected based on the following eligibility criteria: (1) population size between 200–800, (2) within a health facility catchment with an annual incidence of 60–160 cases/1000 population, and (3) an area where the PECADOM+ model was established or eligible for roll-out. To minimize intervention contamination, a buffer of 2.5 km was ensured between village centroids.

### Ethical approval

The study protocol was approved by the Comité National d’Ethique pour la Recherche en Santé (Dakar, Senegal) and the University of California, San Francisco Human Research Protection Program (San Francisco, CA, USA). Stanford investigators only had access to de-identified, aggregated data at the monthly, cluster level.

### Randomization and blinding

Villages were randomized at a 1:1 ratio using a stratified, constrained randomization approach of the following covariates: baseline presence of DSDOMs, health facility, distance to nearest health facility, baseline microscopy-confirmed malaria prevalence, population in 2019, and village population of children <10 years in 2019. An independent trial investigator generated 50 000 randomization schemes and one was randomly sampled from the top 1% (n=500) with the lowest balance scores. Participants, investigators, and the study team were aware of randomized assignment; outcome assessors and laboratory technicians were blinded.

### Interventions

During the intervention year (2021), intervention villages received three rounds of MDA with dihydroartemisinin-piperaquine plus single low-dose primaquine every six weeks. Control villages received three rounds of SMC with sulfadoxine-pyrimethamine plus amodiaquine given to children 3–120 months every four weeks as per standard-of-care. MDA was initiated on June 21, 2021, approximately one month before the presumed start of the transmission season (July– December) and SMC (initiated July 16, 2021) to clear the infectious reservoir (**Figure 1**). The decision to administer MDA every 6 weeks versus 4 weeks was made through discussions with the Tambacounda District Medical Office and to ensure sufficient spacing of drug courses to minimize any potential side effects and based on modeling evidence^7^ suggesting that this interval would provide adequate protection given piperaquine’s long half-life,^28^ and. The timing also ensured comparable coverage of the malaria transmission season relative to SMC. After the three rounds of MDA, study villages were followed up for an additional year (2022), when all villages returned to receiving SMC (**Figure 1**).

### Procedures

Upon selection of eligible villages, the study staff met with administrative, health, and religious leaders of Tambacounda health district to present the trial objectives, describe planned activities, and seek study approval. Local leaders accompanied the study team on visits to each village to explain the study and facilitate community engagement. To raise awareness ahead of each MDA round, the team implemented social media campaigns, radio broadcasts, and television announcements. Town hall meetings and household visits were also conducted to address questions and ensure communities were well informed prior to the campaign.

Before intervention implementation, all study villages received mass distribution of pyrethroid-PBO bednets and a baseline survey was conducted to determine coverage of pyrethroid-PBO nets and malaria prevalence in all ages by microscopy and PCR (**Figure 1**). By March 1, 2021, before the start of the MDA campaign, proactive malaria case management (PECADOM+) was established year-round. At each round, participants were screened for eligibility of receiving study drugs. Individuals were eligible for MDA if they were village residents aged ≥3 months and provided informed consent. Exclusion criteria included self-reported severe or chronic illness, known hypersensitivity to MDA drugs, currently pregnant, recent antimalarial use within the previous three weeks, or reported use of medications affecting cardiac function or prolonging QTc interval within the past four weeks. Participants were ineligible for single low-dose primaquine if breastfeeding or <2 years old. SMC was administered directly by the NMP; who imposed the following eligibility criteria: aged 3-120 months and excluded if they reported acute illness or fever, a known hypersensitivity to SMC drugs, or received antimalarials in the previous 3 weeks or sulfonamide-containing drugs in the previous 10 days. Both SMC and MDA were delivered door-to-door using an age-based dosing strategy (**Appendix Table 6**). All three doses of SMC and MDA drugs were given via directly observed therapy. During drug administration campaigns, suspected cases of malaria were confirmed by RDT and treated with artemether-lumefantrine. Positive cases did not receive chemoprevention until the subsequent cycle.

Data on RDT-confirmed malaria cases were collected throughout the intervention and post-intervention year (2021 and 2022) using health facility and PECADOM+ registries, removing any duplicates. To ensure high-quality malaria case detection, PECADOM+ was scaled-up in all study villages, reaching full implementation by March 1, 2021 and continued until December 31, 2022. Average village population size was estimated by taking the mean across two censuses conducted before and after intervention implementation.

### Outcomes

The primary outcome of the trial was village-level, *P. falciparum*-confirmed malaria incidence during post-intervention transmission season (July–December 2022), defined as the number of RDT-confirmed, symptomatic malaria cases divided by the mean village population size calculated based on two censuses conducted pre- and post-MDA (**Figure 1**). Malaria incidence during the transmission season of the intervention year was considered a secondary outcome.

### Effect measure modifiers

We obtained high-resolution data for all of Senegal on the following potential effect measure modifiers: *Pf* malaria prevalence, precipitation, temperature, vegetation density, % of land covered by surface water, population density, % of population under 10 years, and travel time to nearest health facility. These variables were hypothesized to influence intervention effectiveness through impacting transmission intensity, vector breeding, biting, and survival, and access to care. For example, moderate rainfall can increase vector breeding sites, while prolonged and heavy rainfall may flush mosquito larva from breeding sites, thereby reducing malaria transmission.^29^ Vegetation density, captured by EVI, can increase mosquito breeding and survival by providing refuge and supporting oviposition.^30,31^ Temperature has been shown to have a non-linear influence on *Anopheles* biting rate, vector competence, and survival, and *Pf* parasite development.^32^ Population density has a complex, non-monotonic relationship with malaria; transmission models have linked higher population densities to persistent malaria transmission outside of the peak season.^33^ The proportion of children in a population is another predictor of transmission intensity because children have the highest malaria burden and transmission in this setting.^34^

*Pf* parasite prevalence data, estimated among children 2–10 years of age, in 2020 was obtained from the Malaria Atlas Project at 5m resolution.^35^ Predicted population size per 1 km^2^ was obtained from WorldPop.^36^ For the % of the population under 10 years, in trial sites we used data from the study and in non-trial sites we used data from WorldPop. Daily precipitation data at 0.05° resolution were sourced from the Climate Hazards group Infrared Precipitation with Stations (CHIRPS) dataset.^37^ Using Google Earth Engine, we aggregated these data to calculate total monthly precipitation. Mean monthly temperature data at 0.05° resolution was obtained from the MODIS Monthly CMG Land Surface Temperature and Emissivity (MOD21C3).^38^ While prior studies have shown the importance of accounting for daily temperature variation in predicting malaria transmission,^39^ daily remote sensing datasets at a sufficient spatial resolution had substantial missingness during months with higher rainfall, likely due to cloud cover. Monthly data on the presence of surface water at 30m resolution was sourced from the Joint Research Centre Monthly Water History, v1.4 dataset.^40^ Monthly EVI data at 1 km resolution were obtained from the MODIS/Terra Vegetation Indices Monthly L3 Global 1 km SIN Grid.^41^ Nighttime light radiance (as a proxy for socioeconomic status) were sourced at monthly resolution from the Visible Infrared Imaging Radiometer Suite (VIIRS) Day/Night Band.^42^ Walking and motorized time (in minutes) to nearest health facility were extracted from Malaria Atlas Project at 1 km resolution.^43^ We extracted the mean values of parasite prevalence, population density, % population under 10 years, monthly temperature, and monthly percentage of any surface water for the geocoordinates of each trial village centroid and for all Communes in Senegal. For precipitation, we calculated the monthly minimum, mean, and maximum levels.

To account for potential delayed effects of environmental factors on malaria incidence, we generated lagged variables for precipitation, temperature, and EVI. Specifically, we generated lagged variables of 1 and 2 months for precipitation; 0, 1, and 2 months for temperature; and lags of 0–3 months for EVI. The choice of lags were informed by the following studies: a systematic review of *Plasmodium falciparum* seasonality transmission reported that the most commonly found significant lag in the Sahel region for rainfall was 3 months, while 0–1 month lags were found to be significant for temperature.^44^ Studies of rainfall and malaria in Senegal found associations at 1–2 month lags.^45^ A more recent model of malaria seasonality in Senegal that was tested in other West African countries in the Sahel region found that malaria incidence peaked 1–2 months after the peak of the rainy season for most countries, and incidence peaked 3 months after the rainy season peak in Niger.^46^ A systematic review of *Plasmodium falciparum* transmission seasonality globally found that EVI was associated with transmission from 0–3 months.^44^

Since our intent was to transport estimates to small administrative areas, we chose a temperature dataset that had higher spatial resolution (0.05°) and monthly temporal resolution. A limitation of this approach is that monthly datasets only allowed us to include the mean monthly temperature; minimum and maximum values would have required daily datasets, which had missingness of 10–20%, with higher missingness during the malaria transmission season.

We ultimately excluded surface water, predicted malaria incidence, predicted malaria prevalence, nighttime light radiance, travel time to nearest health facility, and population density from transportability analyses. Predicted surface water was zero in all trial villages and most non-trial Communes, except in a small number of Communes. The distributions of predicted malaria incidence and prevalence, based on Malaria Atlas Project estimates,^35^ were lower in the trial site than in non-trial areas (**Appendix Figure 2**); additionally, MAP estimates had minimal variation across trial clusters and did not correlate with the baseline prevalence measured in the trial clusters (which was assessed by microscopy among all ages), suggesting MAP values did not serve as a strong proxy for overall incidence or prevalence (**Appendix Figure 9**). The range of nighttime light radiance within trial sites was small and did not fully overlap with non-trial sites (−0.09 to 1.0 in trial sites versus 0.04 to 4.2 in non-trial sites). We excluded travel time to health facility because model algorithms did not converge when it was included, resulting in unstable estimates. We excluded population density because it was highly collinear with the percentage of the population under age 10 years, and including age resulted in more stable models.

### Statistical analysis

First, we assessed effect modification of the MDA intervention and by geographic, environmental, and demographic variables. Analyses were conducted at the village-month level across the pre-intervention, intervention, and post-intervention transmission seasons (i.e., July– December of 2020, 2021, and 2022). Indicator variables were used to test for effect modification by Commune. For continuous modifiers, we generated indicators of whether values were above or below median within each follow-up year. To compare incidence rate ratios for MDA vs. control within different levels of potential modifiers, we fit mixed-effects Poisson regression models restricting to each level of the potential effect modifier, which allowed for potential interactions between modifiers and baseline covariates. Models included village-level random intercepts, a log link, an offset for mean village population size during follow-up, and robust standard errors. Consistent with the intention-to-treat analysis performed in the original trial,^4^ malaria case counts were modeled as the dependent variable, and covariates included: trial-year fixed effects, an MDA indicator equal to 1 for random assignment to MDA in the intervention year (2021) and 0 otherwise, a post-MDA indicator equal to 1 for random assignment to MDA in the post-intervention year (2022) and 0 otherwise, an indicator equal to 1 for periods and villages with an existing PECADOM+ model and 0 otherwise to account for differential capture of malaria cases at baseline, and variables included in the constrained randomization (i.e., health post of village, distance to health post, baseline microscopy-confirmed malaria prevalence, village population size, and population size of children <10 years).

To formally test for effect modification on the ratio and additive scale,^19^ we fit models with two-way interaction terms between each modifier and year as well as the indicator for MDA for analyses in the intervention year and the indicator for MDA in the post-intervention year analyses. We defined *R_xz_* as the incidence rate under treatment *X* and modifier *Z*. The ratio scale measure of interaction was defined as (*R*_11_*R*_00_)/(*R*_10_*R*_01_), which can be expressed in terms of incidence ratios (*IR*) as *IR*_11_/(*IR*_10_ *IR*_01_).^19^ We assessed additive scale interaction using the Relative Excess Risk Due to Interaction (RERI), which is defined as *IR*_11_ - *IR*_10_ - *IR*_01_ + 1.^19^ Because MDA had a protective effect on malaria incidence, we recoded the reference levels for the MDA indicator variable and each modifier to that of the lowest risk.^47^ We computed standard errors and confidence intervals using the delta method.

To transport trial estimates to non-trial areas, we applied a doubly robust transportability modeling approach from Dahabreh et al.^20^ We used a non-nested design in which the trial data is concatenated with data from an external target population that partially overlapped with the trial population.^48^ When the difference between the trial and external target populations can be measured by baseline covariates, transportability is similar in principle to direct standardization across multiple covariates. To do so, these methods require data on factors that modify the intervention effect and differ in distribution between trial and target populations.^16^

We restricted our analyses to the subset of Communes where SMC was routinely offered and population density was similar to trial sites (≤152 per 100 m^2^). This was done to minimize potential violations of positivity (i.e., to ensure the trial population could reasonably represent non-trial areas).^15^ Analyses were conducted using a dataset that appended trial and non-trial data, with trial data including treatment assignment, outcome, and effect modifiers, and non-trial data including only effect modifiers. An indicator variable for trial participation (*S_i_*) was included in the combined dataset for each village *i* (for trial data) and Commune *i* (for non-trial data), whereby *S_i_ =*1 for each observation in the trial dataset and *S_i_*=0 for each Commune. Estimates were transported to the Commune level, the smallest standard administrative level in Senegal used for public health planning. We aggregated malaria incidence data by summing the total number of cases and population size and effect modifier data by calculating the minimum, mean, maximum, or total of covariates as described in the prior section.

The doubly robust approach fits a model for the probability of trial participation and a model for the expectation of the outcome—if either is correctly specified, the estimator is consistent.^20^ First, for each Commune, we fit an model for the probability of trial participation Pr(*S_i_*=1|***X****_i,t_*), where *X_i,t_* is a matrix of effect measure modifiers for Commune *i* at month *t* that included variables listed in the previous section. We fit these models using data from trial sites appended with non-trial Communes. We then calculated weights by intervention arm (*A_i_*) in each Commune *i* at month *t* normalized to the sum of the number of non-trial areas as follows:

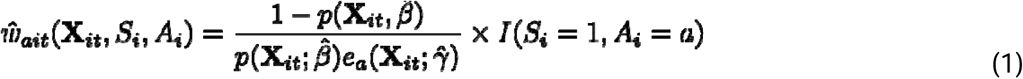

where *A_i_* is an indicator for random assignment to MDA vs. control, 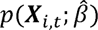 is an estimator of the probability that a Commune would be included in the trial at month *t* (Pr(*S_i_*=1|***X****_i,t_*) and 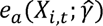 is an estimator of the probability of assignment to treatment arm *a* among trial clusters (Pr(*A*=*a* |***X****_i,t_*,*S_i_* =1)).

Second, we fit outcome models for incident malaria cases in each Commune at month *t* with effect modifiers as independent variables using data from trial sites and stratifying by intervention arm. 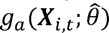 is an estimator for the conditional expectation of the number of malaria cases (E[*Y_i,t_* |***X****_i,t_*, *S_i_* =1, *A_i_*=*a*]), where *Y_i,t_*is the number of malaria cases for each cluster *i* at month *t.* We then predicted outcomes in each arm in non-trial Communes. Outcome models did not adjust for covariates used in the constrained randomization or include the log population offset, as was done in the original trial analysis.^4^ This was because these variables were not available in non-trial areas. However, point estimates and 95% CIs were similar between unadjusted and adjusted analyses in the trial^4^ and when the population offset term was excluded (**Appendix Table 6**). Additionally, we did not adjust for baseline malaria incidence as done in the original trial analysis because we did not have monthly data on malaria cases prior to and during the intervention period in the non-trial areas. A re-analysis comparing the original trial’s estimates restricted to the intervention year (2021) or post-intervention year (2022) found minimal differences in point estimates and 95% CIs in the intervention year and moderate differences in the post-intervention year (**Appendix Table 5**). However, interpretation of findings was consistent between the two models with overlapping 95% CIs.

Given the relatively small number of Communes and potentially large number of covariates, we fit models for the probability of trial participation and for malaria outcomes using elastic net models with ∝=0.5, which used an equal mix of Lasso and Ridge regularization to balance feature selection and handling of correlated predictors. We fit models for 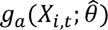 using a quasi-Poisson family and 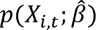 and 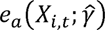 using a binomial family. The relax parameter was set to true, so that models were refit without penalty after the initial variable selection, potentially improving prediction accuracy. To identify the optimal level of regularization, we use 10-fold cross-validation and a lambda sequence of 0.01 to 100, with length 100. We used elastic net model fits with the lambda value that minimized the cross-validated error. If only one covariate was included in the model, we used a generalized linear model with a binomial family instead.

Prior to elastic net model fitting, we performed several forms of covariate screening to mitigate multicollinearity, streamline the covariate set, and improve model stability. First, we excluded any covariates for which there was no overlap in the covariate range in *S*=1 and *S*=0 to minimize empirical violations of the positivity of trial participation assumption. Second, we screened covariates for multicollinearity within predefined variable groups (precipitation, temperature, and EVI) with different lag values. For each group covariates, we calculated pairwise Pearson correlations between variables and estimated the correlation with the outcome. If any correlations were >0.7 within a group of covariates, we retained the covariate from the group with the strongest correlation with the outcome. Third, for models of 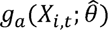, we assessed the association between each covariate and the number of malaria cases using likelihood ratio tests with a Poisson family; we retained covariates for which the p-value was <0.2. We performed a complete case analysis.

We then implemented a doubly robust estimator to estimate intervention effects for each Commune. The estimator uses data from trial clusters as well as non-trial communes. For trial clusters, it uses outcome model predictions as bias correction terms along with observed outcomes, weighted by the inverse probability of selection into the trial. For non-trial Communes in the target population, it relies solely on the outcome model predictions. The double robust estimator 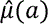 is defined as follows:

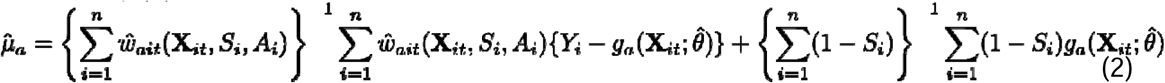

We separately estimated weighted averages for the intervention and control arms, then calculated percent effectiveness as (1 – [incident cases in intervention / incident cases in control]) x 100%. to estimate the transported effect. To obtain 95% CIs, we performed non-parametric bootstrapping by resampling Communes with replacement, with 1000 iterations.

To validate our transportability model, we transported trial estimates in the intervention year (2021) and post-intervention year (2022) to the geographic area surrounding trial villages by creating a convex hull of the trial village centroids (**Appendix Figure 7**). Given that effect modifier values should be highly similar to the geographic areas approximate to the trial site, we expected that this analysis should produce effect estimates close to the original trial. We then extracted effect modifier values from remote sensing data within the trial site, set *S*=0, then re-fit transportability models. We obtained 95% CIs using the bias-corrected and accelerated bootstrap as described above.

To make causal inferences about intervention effectiveness outside the original trial areas, several assumptions are required. First, the assumption of conditional exchangeability of intervention assignment states that clusters assigned to MDA are exchangeable with those assigned to control. Second, the assumption of positivity of intervention assignment states that there is a non-zero probability of intervention assignment in any stratum defined by the covariate distribution of trial clusters. We expect the first two assumptions to hold by randomization. Third, the assumption of conditional exchangeability over trial participation states that the Communes that included trial clusters are exchangeable with those in the target population or with the entire trial conditional on covariates and study arm. Fourth, the assumption of positivity of trial participation states that there is non-zero probability of trial participation in any stratum defined by covariates that are needed to ensure conditional exchangeability. To minimize possible violations of this positivity assumption, we restricted analyses to areas where SMC was offered during the trial period and only proceeded with estimation in Communes where 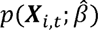 was ≥0.75. Fifth, the consistency assumption states that the potential outcome under each intervention for any cluster is equal to the cluster’s observed outcome, assuming that the intervention is implemented similarly in both trial and target settings. Sixth, the no interference assumption states that a cluster’s potential outcome is not affected by the intervention assignment of other clusters. We expect this assumption to hold based on the ≥2.5 km buffer zone implemented as part of the original trial.^4^

As a separate sensitivity analysis, we investigated whether variation in MDA coverage across trial clusters was associated with effect modifiers included in our transportability models to assess whether effect modifiers were associated with operational feasibility of MDA. We calculated Spearman correlation coefficients between coverage and each effect modifier across intervention clusters.

## Data aailability

The de-identified individual participant data and corresponding data dictionaries may be available upon reasonable request and approval from Jean Louis Ndiaye and Michelle S. Hsiang via a data sharing agreement. A description of study objectives will be needed prior to requests. Any queries for data requests should be made via email to Michelle Roh.

## Code availability

Replication scripts are available at https://github.com/jadebc/senegal-mda-transport-public.

## Acknowledgments

The original cluster-randomized trial was funded by the US President’s Malaria Initiative. Research reported in this publication was supported in part by the Eunice Kennedy Shriver National Institute of Child Health & Human Development of the National Institutes of Health and National Institute of Allergy and Infectious Diseases of the National Institutes of Health under Award Numbers K99HD111572 (PI: MER) and K01AI141616 (PI: JBC). JBC and MSH are Chan Zuckerberg Biohub Investigators. We also acknowledge the Stanford Research Computing Center for computational resources at the Sherlock high-performance cluster. The content is solely the responsibility of the authors and does not necessarily represent the official views of the National Institutes of Health.

## Author contributions

Conceptualization: JBC, MER, MSH, JLN, EKCB

Methodology: JBC, YT, MER, MB

Software: JBC, YT, GBH

Validation: JBC, YT, GBH

Formal analysis: JBC, YT, GBH

Investigation: MER, EBKC, JLN, MSH, AS, AD, ACL

Resources: AF, PM, EBKC, JLN, AS, AD, ACL

Data Curation: MER, YT, GBH

Writing - Original Draft: MER, JBC

Writing - Review & Editing: All authors

Visualization: JBC, GBH

Supervision: JBC

Funding acquisition: JBC

