## Appendix for "Using transportability methods to map the local effectiveness of mass drug administration for malaria in Senegal": NatureMed_MDA Transport_Appendix_FINAL.pdf

**Appendix Table 1. Potential effect modifier values by study arm in July-December 2021**

| | MDA (n=30) | Control (n=30) |
|---|---|---|
| **Precipitation (mm)** | | |
| Minimum (1-month lag) | 109 (93) | 109 (93) |
| Minimum (2-month lag) | 109 (93) | 109 (93) |
| Mean (1-month lag) | 120 (91) | 120 (91) |
| Mean (2-month lag) | 125 (89) | 125 (89) |
| Maximum (1-month lag) | 109 (93) | 109 (93) |
| Maximum (2-month lag) | 109 (93) | 109 (93) |
| **Temperature (°C)** | | |
| Daytime (0-month lag) | 38.4 (3.9) | 38.4 (4.2) |
| Daytime (1-month lag) | 38.6 (4.1) | 38.6 (4.3) |
| Daytime (2-month lag) | 38.6 (4.2) | 38.7 (4.5) |
| Nighttime (0-month lag) | 25.2 (2.7) | 25.2 (2.7) |
| Nighttime (1-month lag) | 25.6 (2.1) | 25.5 (2.1) |
| Nighttime (2-month lag) | 25.0 (2.4) | 25.0 (2.4) |
| **Enhanced vegetation index** | | |
| 1-month lag | 3683 (1338) | 3700 (1361) |
| 2-month lag | 3675 (1354) | 3682 (1380) |
| **Other** | | |
| Population density (per 3 arc-seconds) | 36.9 (33.2) | 50.4 (39.5) |
| % of population aged < 10 years | 38.1 (4.4) | 37.2 (5.0) |
| Nighttime light radiance | 0.3 (0.1) | 0.3 (0.1) |
| **Travel time to nearest health care facility (minutes)** | | |
| Walking time | 183.0 (112.0) | 179.5 (111.8) |
| Motorized time | 29.0 (21.6) | 29.7 (22.0) |

Columns report the mean (SD) of each variable.

**Appendix Table 2. Percent reduction in malaria incidence for MDA vs. control stratified by potential effect modifiers**

| Effect modifiers | 2021 | 2022 |
|---|---|---|
| **Commune** | | |
| Koussanar | 30% (21%, 60%) | -3% (-88%, 43%) |
| Sinthiou Maleme | 63% (-31%, 89%) | 44% (-142%, 87%) |
| Netteboulou | 70% (58%, 79%) | -14% (-62%, 20%) |
| Missirah | 60% (-10%, 86%) | 54% (21%, 73%) |
| **Temperature** | | |
| *Nighttime mean* | | |
| Below median | 59% (18%, 80%) | 17% (-56%, 56%) |
| Above median | 41% (-4%, 67%) | 33% (-24%, 64%) |
| *Daytime mean* | | |
| Below median | 64% (34%, 81%) | 24% (-37%, 58%) |
| Above median | 42% (8%, 63%) | 5% (-57%, 42%) |
| **Precipitation** | | |
| *Minimum* | | |
| Below median | 58% (32%, 74%) | 40% (-1%, 64%) |
| Above median | 65% (35%, 82%) | 18% (-55%, 57%) |
| *Mean* | | |
| Below median | 56% (31%, 72%) | 33% (-8%, 59%) |
| Above median | 60% (21%, 79%) | 11% (-65%, 52%) |
| *Maximum* | | |
| Below median | 58% (32%, 74%) | 40% (-1%, 64%) |
| Above median | 65% (35%, 82%) | 18% (-55%, 57%) |
| **Population density** | | |
| Below median | 26% (-31%, 58%) | -22% (-127%, 34%) |
| Above median | 69% (43%, 83%) | 50% (10%, 72%) |
| **% population < 10 years** | | |
| Below median | 64% (33%, 81%) | 57% (25, 75%) |
| Above median | 39% (-12%, 67%) | -22% (-129%, 34%) |
| **Health facility walk time** | | |
| Below median | 50% (7%, 73%) | 21% (-44%, 57%) |
| Above median | 61% (25%, 79%) | 34% (-27%, 66%) |
| **Health facility motorized time** | | |
| Below median | 50% (10%, 72%) | 25% (-34%, 58%) |
| Above median | 59% (22%, 78%) | 34% (-29%, 66%) |
| **Enhanced vegetation index** | | |
| Below median | 56% (27%, 74%) | 33% (-6%, 58%) |
| Above median | 35% (-18%, 64%) | 3% (-86%, 50%) |
| **Nighttime light radiance** | | |
| Below median | 30% (-21%, 60%) | -3% (-88%, 43%) |
| Above median | 63% (-31%, 89%) | 44% (-142%, 87%) |

**Appendix Table 3. Heterogeneity of the effect of MDA vs. control in the intervention year (2021)**

| Potential modifier | Observed joint incidence ratio for MDA and modifier | Expected joint incidence ratio for MDA and modifier | Measure of effect modification on ratio scale | Ratio scale p-value | Measure of effect modification on additive scale (95% CI) | Additive scale p-value |
|---|---|---|---|---|---|---|
| **Commune** | | | | | | |
| Koussanar | 1.88 | 0.36 | 5.29 | 0.105 | -3.29 | 0.057 |
| Sinthiou Maleme | 0.70 | 0.45 | 1.57 | 0.659 | -2.76 | 0.510 |
| Netteboulou | 1.12 | 0.47 | 2.39 | 0.706 | -1.54 | 0.231 |
| Missirah | 0.74 | 0.62 | 1.19 | 0.556 | -0.45 | 0.737 |
| **Temperature** | | | | | | |
| Nighttime mean | 1.05 | 0.48 | 2.21 | 0.82 | -1.45 | 0.107 |
| Daytime mean | 1.34 | 0.45 | 2.98 | 0.31 | -2.19 | 0.072 |
| **Precipitation** | | | | | | |
| Minimum | 1.43 | 0.29 | 4.88 | 0.203 | -0.32 | 0.768 |
| Mean | 1.03 | 0.47 | 2.18 | 0.903 | -1.39 | 0.096 |
| Maximum | 1.43 | 0.29 | 4.88 | 0.203 | -0.32 | 0.768 |
| **Other** | | | | | | |
| % population age <10 years | 2.24 | 0.24 | 9.32 | 0.048 | 1.22 | 0.421 |
| Population density | 0.40 | 0.86 | 0.47 | 0.020 | 1.73 | 0.235 |
| Enhanced vegetation index | 1.34 | 0.34 | 3.95 | 0.236 | -0.42 | 0.565 |
| Nighttime light radius | 1.15 | 0.45 | 2.58 | 0.563 | -1.68 | 0.107 |
| Motorized time to healthcare facility | 0.88 | 0.55 | 1.60 | 0.764 | -0.89 | 0.520 |
| Walk time to healthcare facility | 0.83 | 0.49 | 1.70 | 0.662 | -1.81 | 0.260 |

For communes, incidence ratios compare incidence in the MDA arm in a given commune to that in the control arm in all other communes. Incidence ratios were adjusted for timing of case detection relative to full PECADOM+ scale-up, similar to the original trial analysis. Other effect modifiers were coded as 1 for clusters with monthly values above in 2021 and 0 for values below the median. The observed joint incidence ratio compared the malaria incidence for MDA and modifier level 1 vs. control and modifier level 0. The expected joint incidence ratio = (incidence ratio for MDA and modifier level 0 vs. control and modifier level 0) x (incidence ratio for control and modifier level 1 vs. control and modifier level 0). The measure of effect modification on multiplicative scale is the observed joint incidence ratio divided by the expected joint incidence ratio. The measure of effect modification on additive scale is the relative excess risk due to interaction. p-values were obtained by the delta method.

**Appendix Table 4. Heterogeneity of the effect of MDA vs. control in the post-intervention year (2022)**

| Potential modifier | Observed joint incidence ratio for MDA and modifier | Expected joint incidence ratio for MDA and modifier | Measure of effect modification on ratio scale | Ratio scale p-value | Measure of effect modification on additive scale (95% CI) | Additive scale p-value |
|---|---|---|---|---|---|---|
| **Commune** | | | | | | |
| Koussanar | 1.32 | 1.47 | 0.90 | 0.404 | -0.13 | 0.868 |
| Sinthiou Maleme | 0.75 | 1.68 | 0.45 | 0.603 | -0.07 | 0.953 |
| Netteboulou | 1.98 | 1.55 | 1.28 | 0.013 | 0.46 | 0.497 |
| Missirah | 0.64 | 2.09 | 0.31 | 0.036 | -1.31 | 0.005 |
| **Temperature** | | | | | | |
| Nighttime mean | 0.72 | 2.05 | 0.35 | 0.271 | -1.24 | 0.059 |
| Daytime mean | 0.94 | 1.89 | 0.50 | 0.861 | -0.85 | 0.257 |
| **Precipitation** | | | | | | |
| Minimum | 1.69 | 0.95 | 1.79 | 0.099 | -1.78 | 0.003 |
| Mean | 1.41 | 1.42 | 0.99 | 0.352 | 0.01 | 0.995 |
| Maximum | 1.69 | 0.95 | 1.79 | 0.099 | -1.78 | 0.003 |
| **Other** | | | | | | |
| % population age <10 years | 2.31 | 0.80 | 2.88 | 0.008 | -2.27 | 0.001 |
| Population density | 0.56 | 2.51 | 0.22 | 0.091 | -1.80 | 0.008 |
| Enhanced vegetation index | 1.31 | 1.05 | 1.24 | 0.440 | -1.43 | 0.045 |
| Nighttime lights | 1.50 | 1.33 | 1.13 | 0.226 | 0.18 | 0.775 |
| Motorized time to healthcare facility | 0.97 | 1.76 | 0.55 | 0.939 | -0.71 | 0.286 |
| Walk time to healthcare facility | 0.93 | 1.66 | 0.56 | 0.832 | -0.50 | 0.450 |

For communes, incidence ratios compare incidence in the MDA arm in a given commune to that in the control arm in all other communes. Incidence ratios were adjusted for timing of case detection relative to full PECADOM+ scale-up, similar to the original trial analysis. Other effect modifiers were coded as 1 for clusters with monthly values above in 2022 and 0 for values below the median. The observed joint incidence ratio compared the malaria incidence for MDA and modifier level 1 vs. control and modifier level 0. The expected joint incidence ratio = (incidence ratio for MDA and modifier level 0 vs. control and modifier level 0) x (incidence ratio for control and modifier level 1 vs. control and modifier level 0). The measure of effect modification on multiplicative scale is the observed joint incidence ratio divided by the expected joint incidence ratio. The measure of effect modification on additive scale is the relative excess risk due to interaction. p-values were obtained by the delta method.

**Appendix Figure 1. Temporal variation of time-varying effect modifiers during trial follow-up.**

Includes data from communes included in the transportability analysis: N=122 in the intervention year (2021) and N=119 in the post-intervention year (2022). Colored lines indicate means within each commune, and black lines means within the trial clusters.

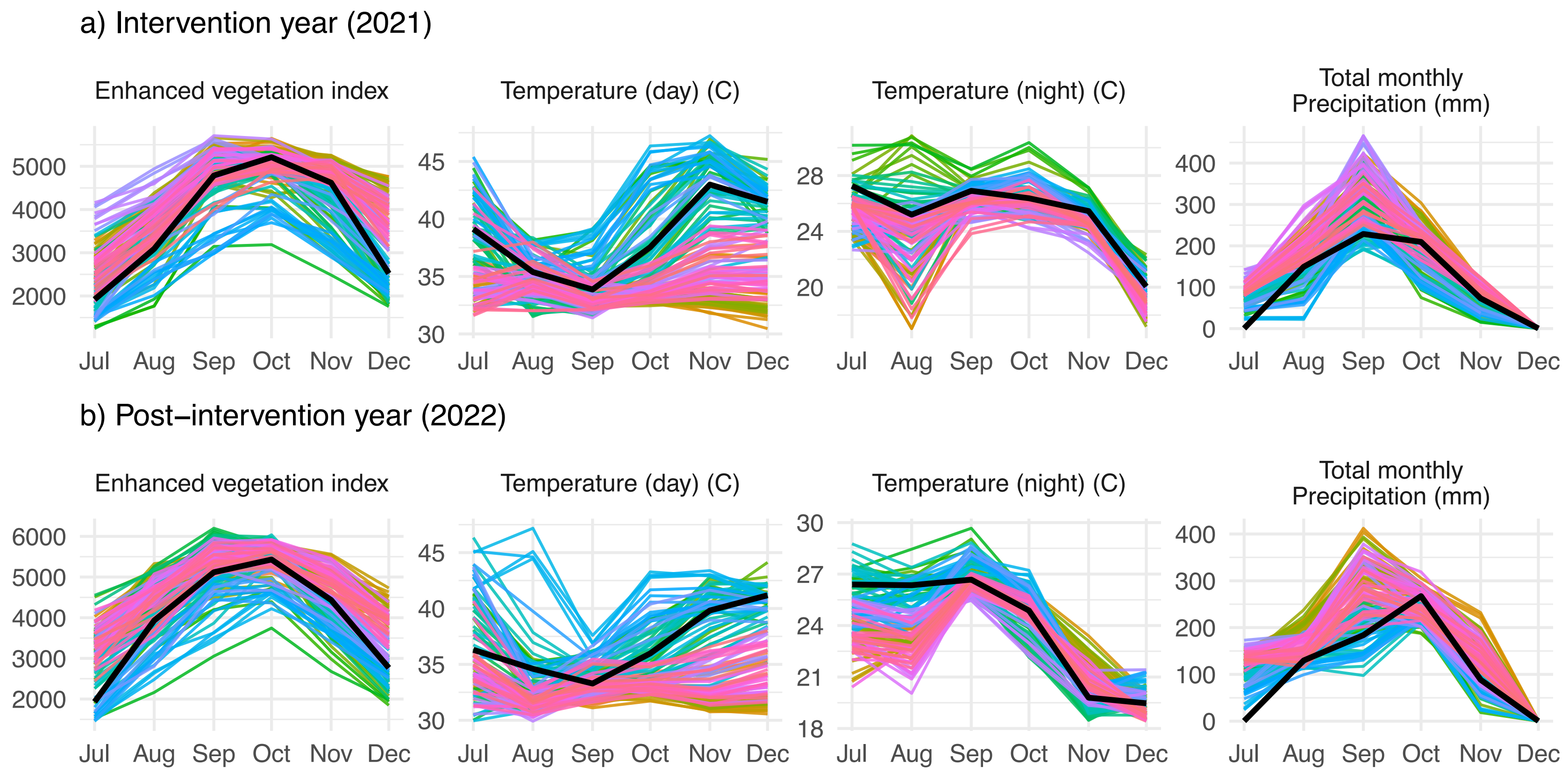

**Appendix Figure 2. Characteristics of communes not included in the transportability analysis.**

This subset of communes includes those with a population size >152 per km, with a predicted probability of trial participation ≤0.75, or where SMC was not offered during the trial period. The color of each point indicates the standardized mean difference for a given covariate in a given commune, calculated as the mean in a non-trial commune minus the mean in the trial site divided by the pooled standard deviation. Communes are sorted from north to south.

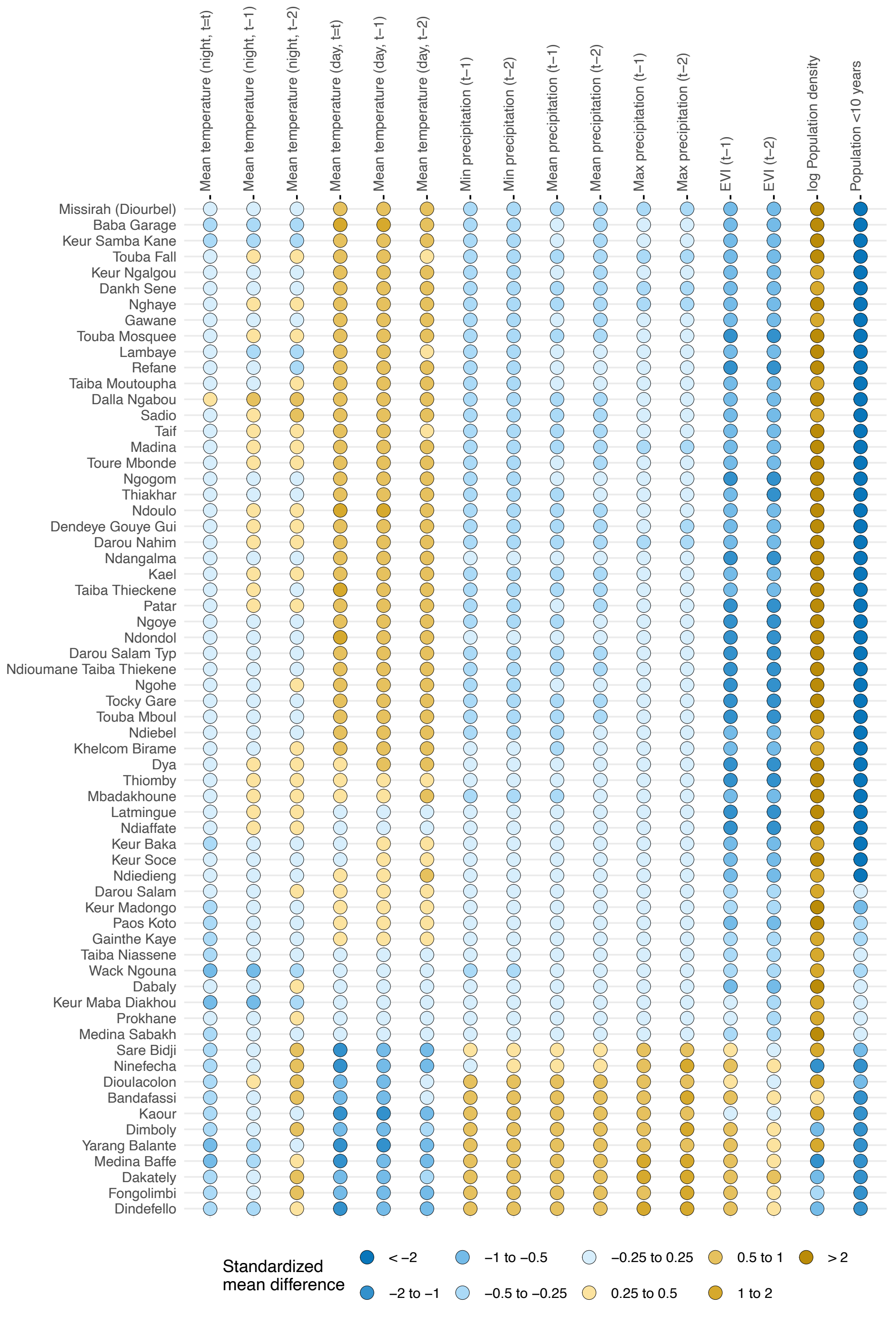

**Appendix Figure 3. Covariate distribution of effect modifiers of Communes within and outside the trial site included in the transportability analysis.**

Data for trial villages are shown in orange, and data for Communes included in the transportability analysis are shown in blue. The number of observations is shown as village-months in the trial site and as Commune-months outside the trial site.

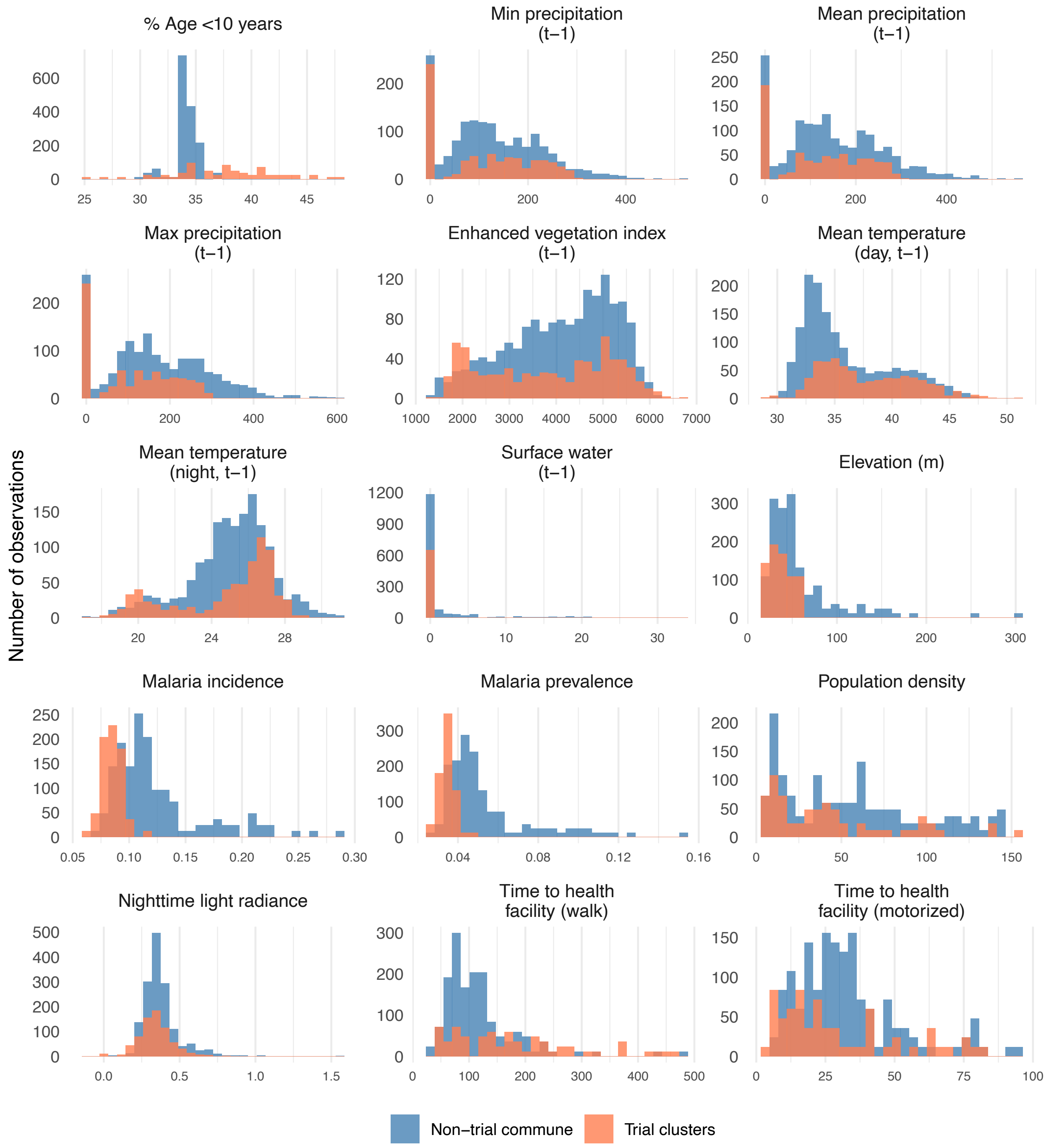

### Appendix Figure 4. Predicted probabilities of trial participation for each commune included in the transportability analysis.

Predicted probabilities P(S) were obtained from elastic net regression models for analyses in which the P(S)>0.75. Analyses were performed separately for each commune using monthly data from July to December (N=366 per commune). Data for trial villages are shown in orange, and data for Communes included in the transportability analysis are shown in blue. Covariates considered for trial participation models included precipitation, temperature, enhanced vegetation index, and population density. The final covariate list used in models varied between communes following screening for collinearity, data sparsity, and feature selection using elastic net regression. Analyses restricted to communes with population size ≤152 per km and where SMC was offered during the trial period. Common support = the percentage of months in a given non-trial Commune for which P(S) was within the range of the study clusters' P(S) for that analysis.

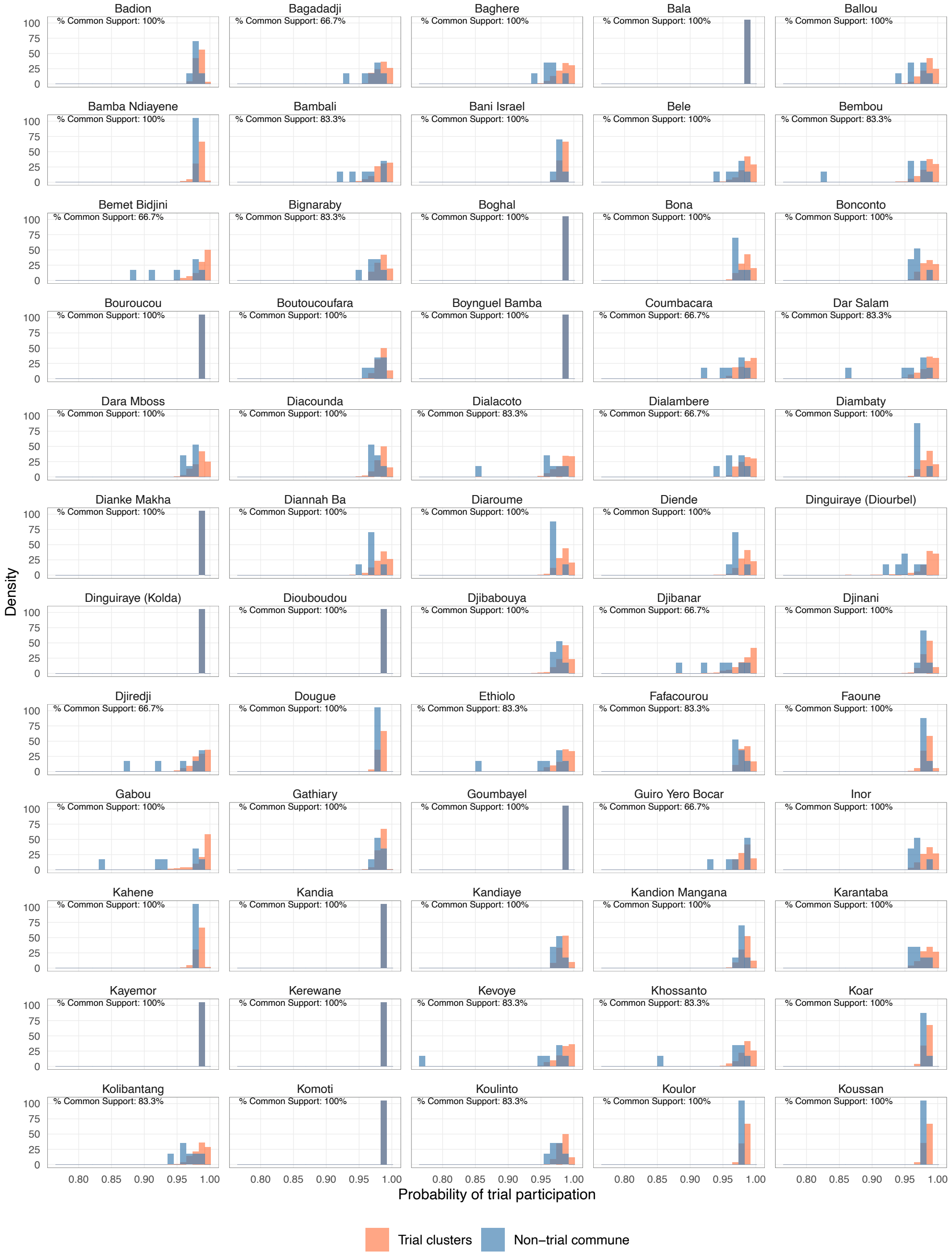

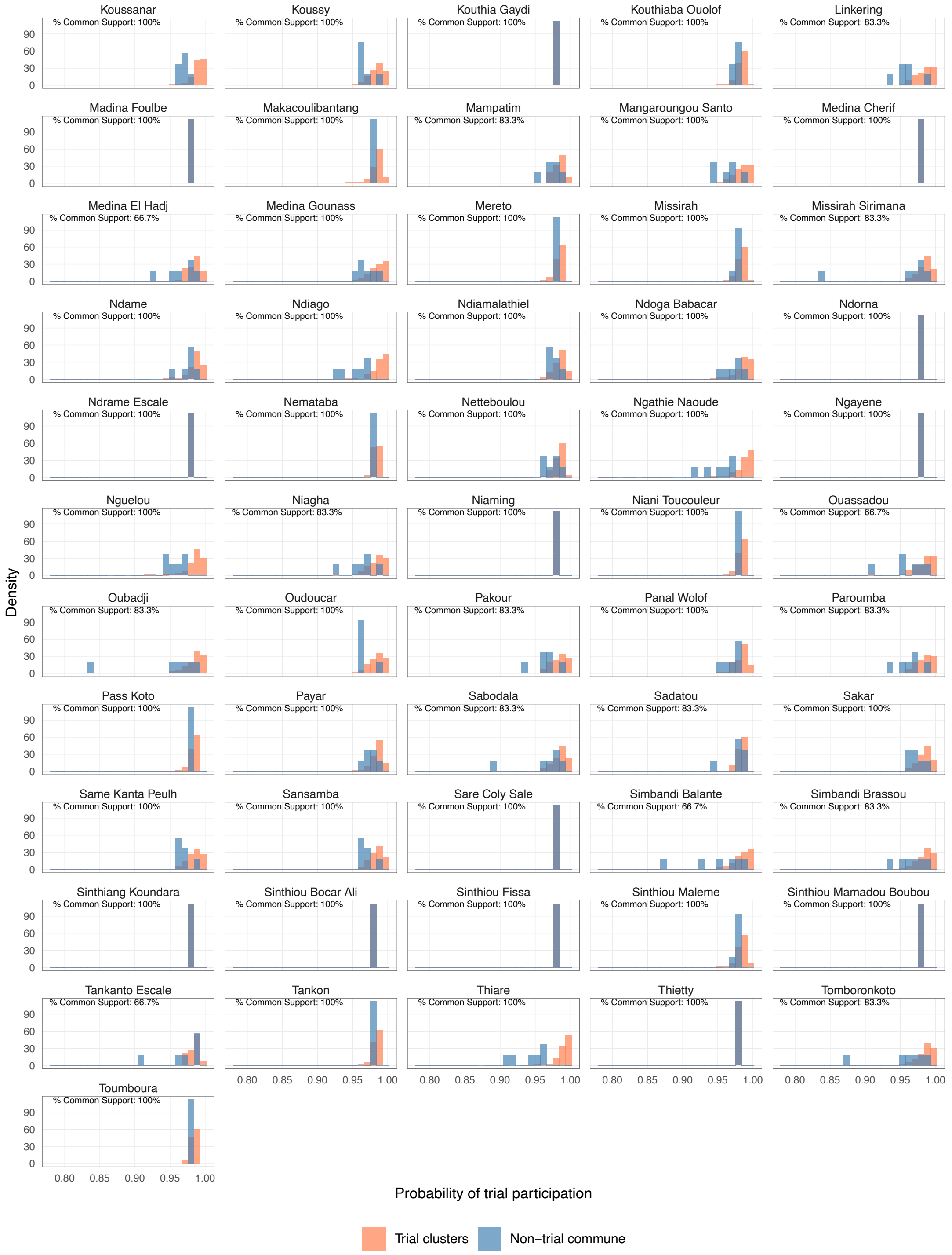
Koussanar
% Common Support: 100%
Koussy
% Common Support: 100%
Kouthia Gaydi
% Common Support: 100%
Kouthiaba Ouolof
% Common Support: 100%
Linkering
% Common Support: 83.3%
Madina Foulbe
% Common Support: 100%
Makacoulibantang
% Common Support: 100%
Mampatim
% Common Support: 83.3%
Mangaroungou Santo
% Common Support: 100%
Medina Cherif
% Common Support: 100%
Medina El Hadj
% Common Support: 66.7%
Medina Gounass
% Common Support: 100%
Mereto
% Common Support: 100%
Missirah
% Common Support: 100%
Missirah Sirimana
% Common Support: 83.3%
Ndame
% Common Support: 100%
Ndiago
% Common Support: 100%
Ndiamalathiel
% Common Support: 100%
Ndoga Babacar
% Common Support: 100%
Ndorna
% Common Support: 100%
Ndrame Escale
% Common Support: 100%
Nemataba
% Common Support: 100%
Netteboulou
% Common Support: 100%
Ngathie Naoude
% Common Support: 100%
Ngayene
% Common Support: 100%
Nguelou
% Common Support: 100%
Niagha
% Common Support: 83.3%
Niaming
% Common Support: 100%
Niani Toucouleur
% Common Support: 100%
Ouassadou
% Common Support: 66.7%
Oubadji
% Common Support: 83.3%
Oudoucar
% Common Support: 100%
Pakour
% Common Support: 83.3%
Panal Wolof
% Common Support: 100%
Paroumba
% Common Support: 83.3%
Pass Koto
% Common Support: 100%
Payar
% Common Support: 100%
Sabodala
% Common Support: 83.3%
Sadatou
% Common Support: 83.3%
Sakar
% Common Support: 100%
Same Kanta Peulh
% Common Support: 100%
Sansamba
% Common Support: 100%
Sare Coly Sale
% Common Support: 100%
Simbandi Balante
% Common Support: 66.7%
Simbandi Brassou
% Common Support: 83.3%
Sinthiang Koundara
% Common Support: 100%
Sinthiou Bocar Ali
% Common Support: 100%
Sinthiou Fissa
% Common Support: 100%
Sinthiou Maleme
% Common Support: 100%
Sinthiou Mamadou Boubou
% Common Support: 100%
Tankanto Escale
% Common Support: 66.7%
Tankon
% Common Support: 100%
Thiare
% Common Support: 100%
Thietty
% Common Support: 100%
Tomboronkoto
% Common Support: 83.3%
Toumboura
% Common Support: 100%
Density
0 30 60 90
0.80 0.85 0.90 0.95 1.00
Probability of trial participation
Trial clusters
Non−trial commune

**Appendix Table 5. Sensitivity analysis of original trial analysis to inclusion of pre-intervention year in the model**

| | Intervention year (2021) | | Post-intervention year (2022) | |
|---|---|---|---|---|
| | Pre-intervention | No pre-intervention | Pre-intervention | No pre-intervention |
| Unadjusted | 55% (28%, 71%) | 47% (23%, 63%) | 26% (-18%, 53%) | 15% (-16%, 37%) |
| Adjusted* | 55% (28%, 71%) | 49% (25%, 66%) | 26% (-17%, 53%) | 4% (-28%, 29%) |

Table cells show intervention effectiveness, defined as (1 - IRR for MDA vs. control) x 100%, where IRR is the malaria incidence rate ratio. 95% confidence intervals are shown in parentheses.

* Adjusted for trial-year fixed effects (minus 2020 baseline data for 'no pre-intervention' analyses), an indicator variable equal to 1 for periods and villages with an existing PECADOM model and 0 otherwise to account for differential capture of malaria cases at baseline, and variables included in the constrained randomization (i.e., health post of village, distance to health post, baseline microscopy-confirmed malaria prevalence, village population size, and population size of children <10 years)

**Appendix Table 6. Sensitivity analysis of original trial analysis to covariate adjustment and log population offset**

| | Intervention year (2021) | | Post-intervention year (2022) | |
|---|---|---|---|---|
| | Offset | No offset | Offset | No offset |
| Unadjusted in original analysis* | 55% (28%, 71%) | 55% (27%, 72%) | 26% (-18%, 53%) | 26% (-20%, 55%) |
| Fully unadjusted | 52% (21%, 71%) | 52% (21%, 71%) | 21% (-20%, 54%) | 21% (-26%, 51%) |
| Adjusted† | 55% (28%, 71%) | 55% (28%, 71%) | 26% (-17%, 53%) | 26% (-18%, 55%) |

Table cells show intervention effectiveness, defined as (1 - IRR for MDA vs. control) x 100%, where IRR is the malaria incidence rate ratio. 95% confidence intervals are shown in parentheses.

* Included an indicator equal to 1 in periods when proactive community case management of fever occurred in the village and 0 otherwise to account for differential capture of malaria cases at baseline.

† Adjusted for trial-year fixed effects, an indicator variable for periods and villages with an existing PECADOM model, and variables included in the constrained randomization (i.e., health post of village, distance to health post, baseline microscopy-confirmed malaria prevalence, village population size, and population size of children <10 years)

**Appendix Figure 5. Spatial distribution of transported effects of mass drug administration in the post-intervention year (2022).**

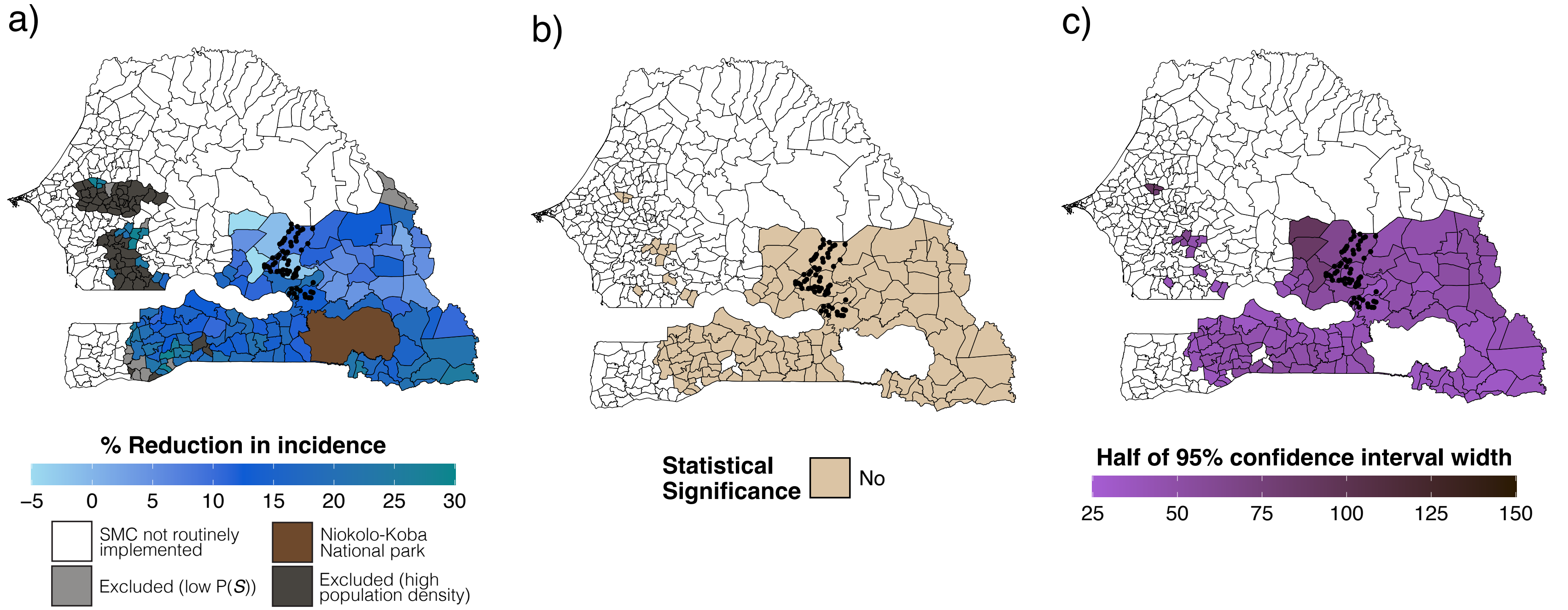

Polygons indicate communes. Panel (a) shows the percent reduction in malaria, expressed as 1 - the ratio of cases in the MDA vs. control arm during the transmission season of the post-intervention year (2022), estimated using doubly robust transportability models. Covariates included in transportability models included precipitation, temperature, enhanced vegetation index, and population density. The final covariate included in transportability models varied between communes following screening for collinearity, data sparsity, association with the malaria case count, and feature selection using elastic net regression. Transportability analyses were restricted to communes where SMC was routinely offered during the trial period, had a population size <152 per km, and a predicted probability of trial selection (P(*S*)) of >0.75. Separate analyses were performed for each Commune-month (N=366 per Commune). Panel (b) indicates whether the 95% confidence interval for transported estimates for each commune included the null. Panel (c) depicts uncertainty, shown as half the length of the 95% confidence interval, estimated using a non-parametric bootstrapping procedure that resampled commune-months within trial datasets and months within the non-trial commune 1,000 times with replacement.

**Appendix Figure 6. Transported effect estimates of mass drug administration to non-trial areas during the post-intervention year (2022) and covariates used in transportability analysis.**

Panel (a) depicts covariates used in the transportability analysis for each commune. Colored points indicate covariates included in the transportability models after screening for collinearity, data sparsity, association with the malaria case count, and feature selection using elastic net regression. Blue and yellow shaded points indicate the standardized mean difference for a given covariate in a given commune, calculated as the mean difference between the non-trial commune and trial site divided by the pooled standard deviation. White points indicate covariates that did not pass screening. In Panel (b), colored points indicate transported effect for each Commune, expressed as the percent reduction in malaria incidence (1 – the ratio of cases in the MDA vs. control arm) in the post-intervention year (2022). Colored vertical lines indicate 95% confidence interval for each commune, which was obtained from non-parametric bootstrap that resampled commune-months within trial datasets and months within non-trial areas for a given commune 1,000 times with replacement. Transportability analyses were performed separately for each commune using monthly data from July to December (N=366 per commune). Transportability analyses restricted to communes with population size ≤152 per km, a predicted probability of trial participation >0.75, and where SMC was offered during the trial period.

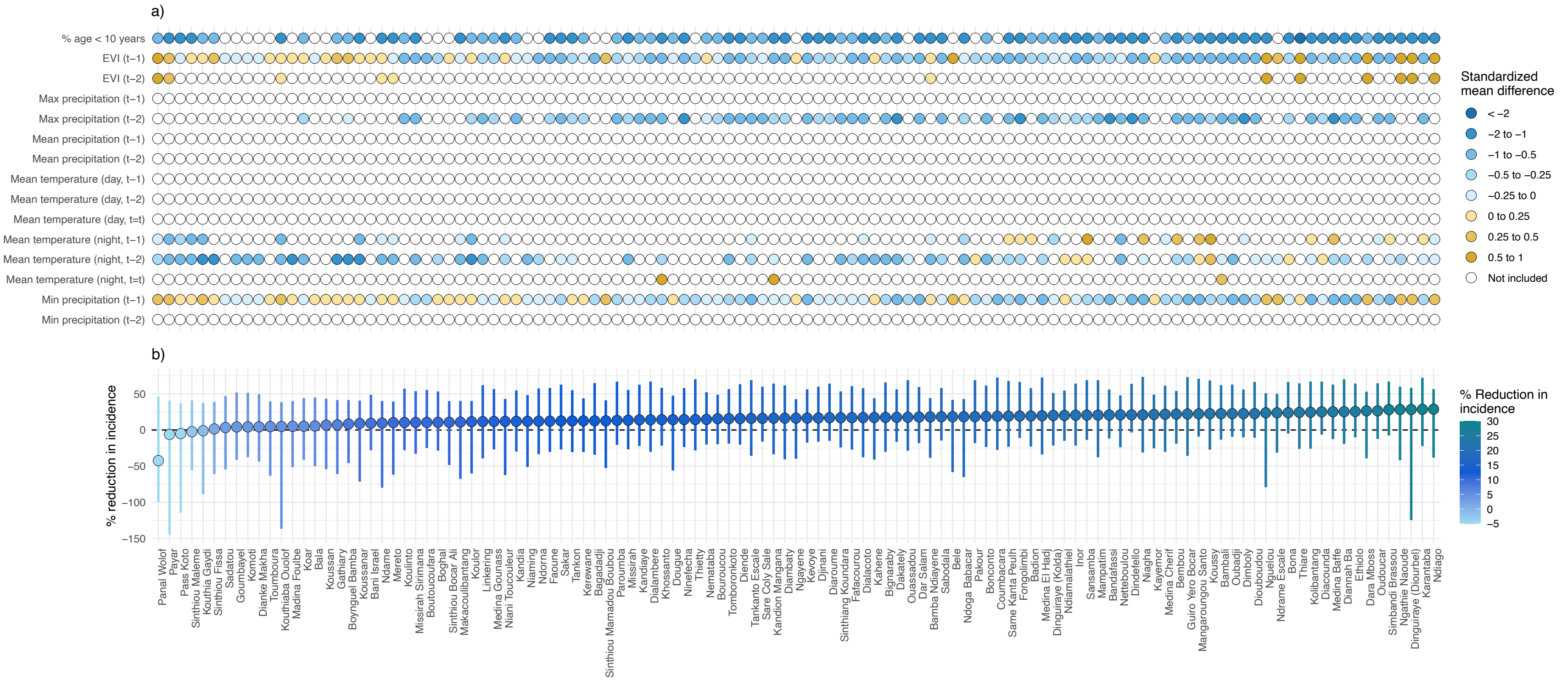

**Appendix Figure 7. Geographic area surrounding trial villages used to validate transportability analyses.**

To validate our transportability model, we transported trial estimates to the area surrounding the trial villages (highlighted in green), approximated by a convex hull of trial village centroids. Because effect modifier values in this area were expected to closely resemble those in the trial villages, transported estimates were anticipated to align with original trial results.

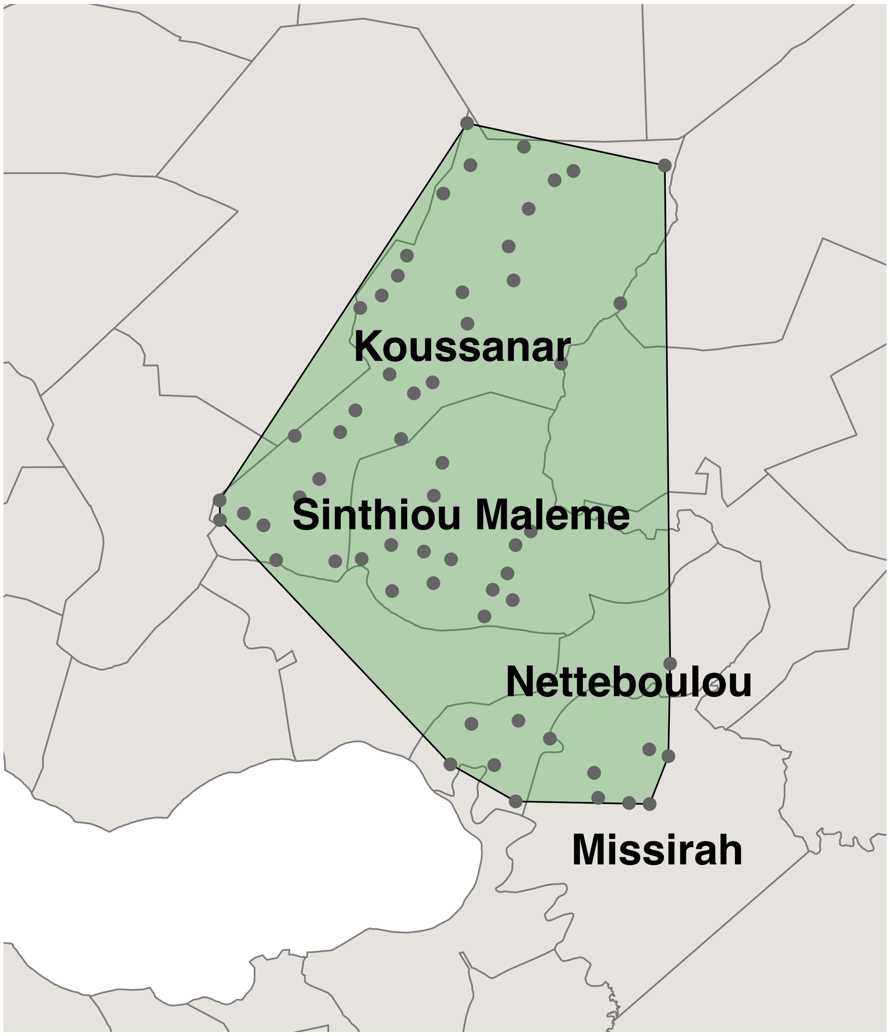

### Appendix Figure 8. Correlation between MDA coverage and effect modifiers during the intervention year (2021).

In the original trial, coverage of MDA varied from 54–80% across intervention clusters. As such, we expect that the variation in transported estimates may also be partially explained by differences in coverage between trial clusters in addition to variation in demographic and environmental settings. To evaluate this, we calculated Spearman's correlation coefficients between MDA coverage and each effect modifier, exclusively among intervention clusters. Y-axis represents the median village-level coverage of all three doses of dihydroartemisinin-piperaquine, averaged across the three rounds. Effect modifier values were calculated by extracting effect modifier values from each village; for time-varying values (e.g., precipitation, temperature, and EVI), we took the mean values across the 2021 transmission season (July-December). As shown in the plots, correlations were generally weak (|ρ| < 0.2) for most modifiers, except for daytime temperature, precipitation, and travel time to nearest health facility.

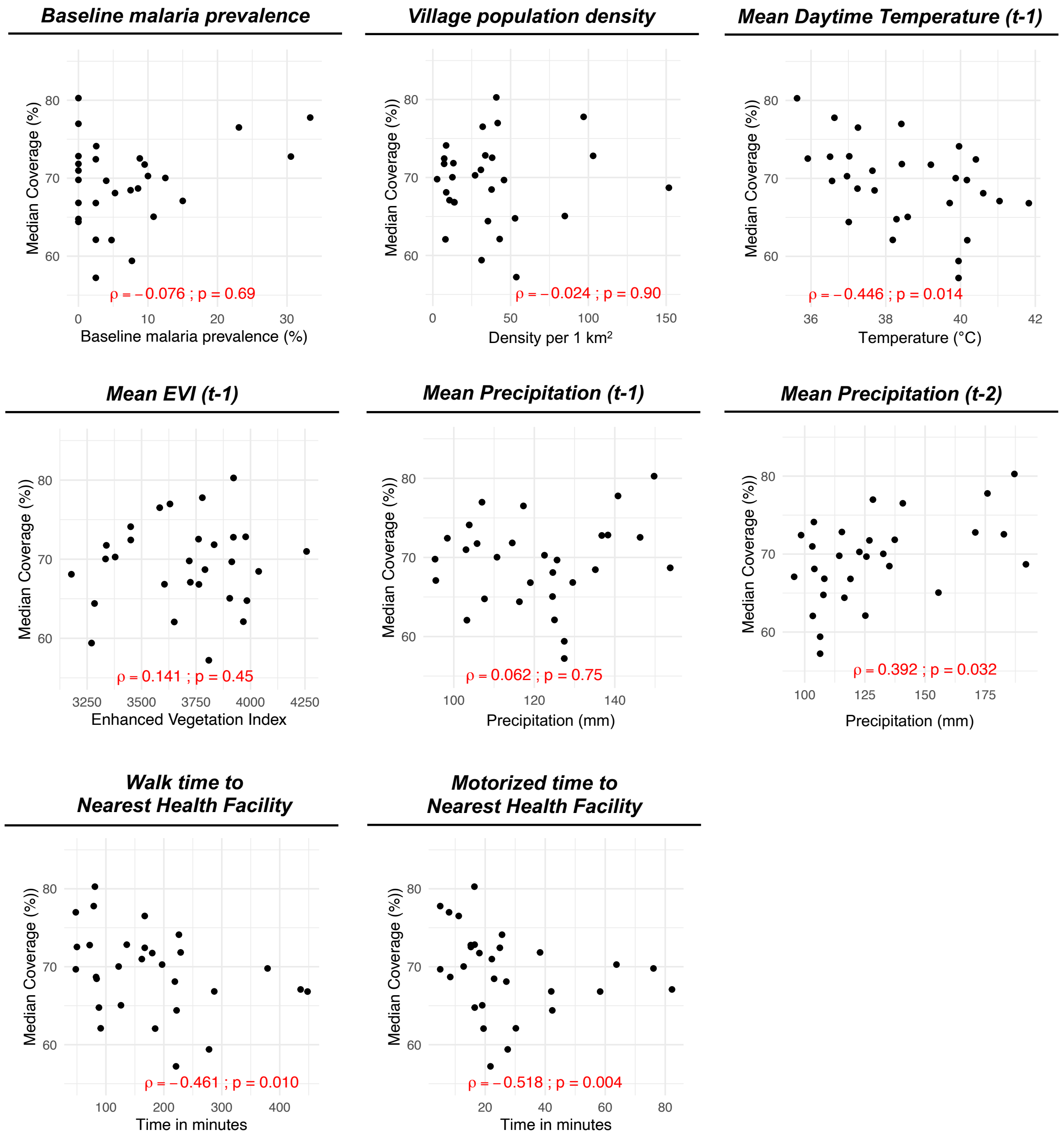

**Appendix Table 7. Dosing regimens for dihydroartemisinin-piperaquine, single-low dose primaquine, and sulfadoxine-pyrimethamine + amodiaquine**

**Mass drug administration (intervention arm):**

| **Dihydroartemisinin-piperaquine** | | | |
|---|---|---|---|
| Age | Number of tablets containing 40 mg dihydroartemisinin and 320 mg piperaquine | | |
| | Day 1 | Day 2 | Day 3 |
| 3 to 24 months | 0.5 | 0.5 | 0.5 |
| 2 to 7 years | 1 | 1 | 1 |
| 8 to 10 years | 1.5 | 1.5 | 1.5 |
| 11 to 14 years | 2 | 2 | 2 |
| 15+ years | 3 | 3 | 3 |
| **Primaquine (given on day 1)** | | | |
| Age | Number of tablets | Dosage (mg) | Volume (ml) of water used to dilute tablets |
| 2 to 4 years | 0.5 | 3.75 | 3 |
| 5 to 7 years | 0.75 | 5.625 | 5 |
| 8 to 10 years | 1 | 7.5 | 10 |
| 11 to 13 years | 1.5 | 11.25 | -- |
| 14+ years | 2 | 15 | -- |

**Seasonal malaria chemoprevention (control arm):**

| **Sulfadoxine-pyrimethamine + amodiaquine (SP-AQ)** | | | |
|---|---|---|---|
| Age | Number of tablets containing 500 mg sulfadoxine and 25 mg pyrimethamine, and 150 mg amodiaquine | | |
| | Day 1 | Day 2 | Day 3 |
| 3 to 11 months | 0.5 tablet SP + 0·5 tablet AQ | 0.5 tablet AQ | 0.5 tablet AQ |
| 12 to 59 months | 1 tablet SP + 1 tablet AQ | 1 tablet AQ | 1 tablet AQ |
| 60 to 120 months | 1.5 tablets SP + 1.5 tablets AQ | 1.5 tablets AQ | 1.5 tablets AQ |

**Appendix Figure 9. Comparison of observed baseline malaria prevalence vs. predicted malaria prevalence.**

In the original trial, baseline *Plasmodium falciparum* malaria prevalence was assessed by microscopy in all ages at the end of the transmission season (December 10-20, 2020). Malaria Atlas Project predicts *Plasmodium falciparum* malaria prevalence at fine spatial scales among children 2-10 years using a combination of rapid diagnostic test- and microscopy-based surveys.

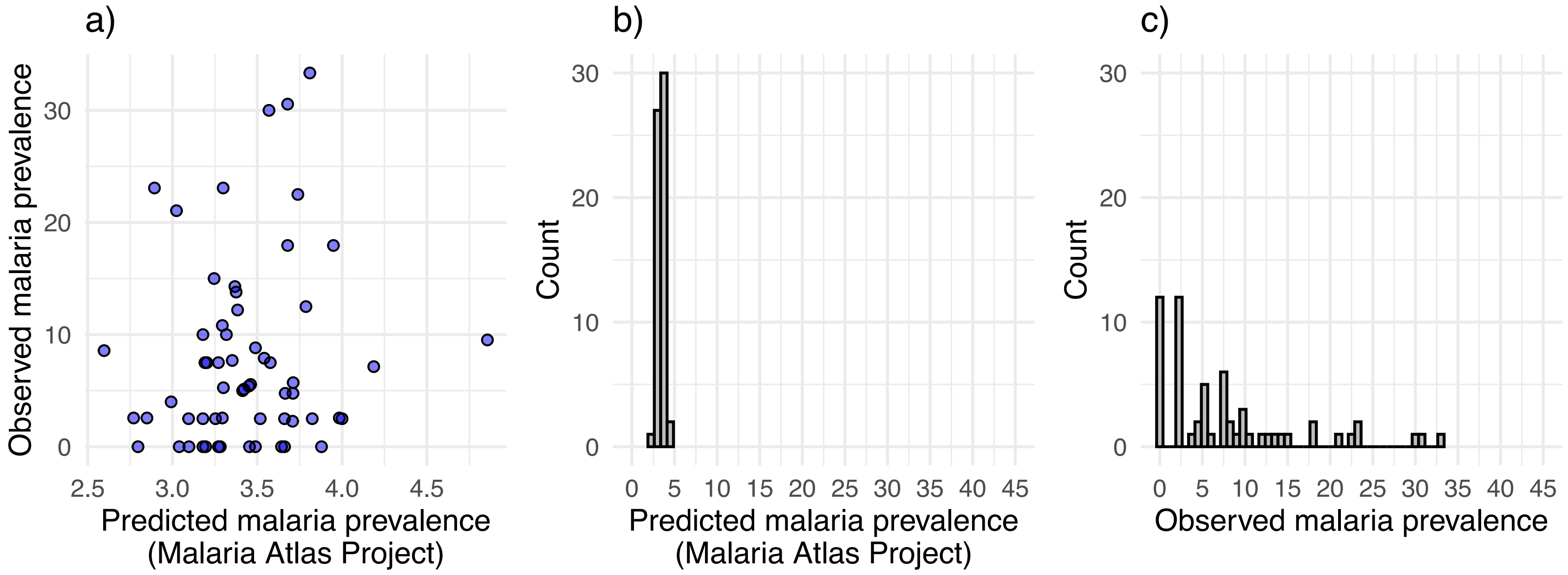
